# Automated stenosis estimation of coronary angiographies using end-to-end learning

**DOI:** 10.1101/2024.10.08.24315070

**Authors:** Christian Kim Eschen, Karina Banasik, Anders Bjorholm Dahl, Piotr Jaroslaw Chmura, Peter Bruun-Rasmussen, Frants Pedersen, Lars Køber, Thomas Engstrøm, Morten Bøttcher, Simon Winther, Alex Hørby Christensen, Henning Bundgaard, Søren Brunak

## Abstract

**Background:** The initial evaluation of coronary stenosis during coronary angiography is typically performed by visual assessment. The visual assessment of coronary angiographies has limited accuracy compared to quantitative methods like fractional flow reserve and quantitative coronary angiography. Quantitative methods are also more time-consuming and costly.

**Objectives:** To test whether applying deep-learning-based image analysis to coronary angiographies might yield a faster and more accurate stenosis estimation than visual assessment.

**Methods:** We developed deep learning models for predicting coronary artery stenosis using 332,582 multi-frame x-ray images (cine loops) from 19,414 patients undergoing coronary angiography. The curated dataset for model development included 13,840 patients, with 62,165 cine loops of the left coronary artery and 31,161 cine loops of the right coronary artery.

**Results:** For identification of significant coronary stenosis (visual assessment of diameter stenosis >70%), our model obtained a receiver operator characteristic (ROC) area under the curve (ROC-AUC) of 0.903 (95% CI: 0.900-0.906) on the internal test set with 5,056 patients. The performance was evaluated on an external test set with 608 patients against visual assessment, 3D quantitative coronary angiography, and fractional flow reserve (≤ 0.80), obtaining ROC AUC values of 0.833 (95% CI: 0.814-0.852), 0.798 (95% CI: 0.741-0.842, and 0.780 (95% CI: 0.743-0.817), respectively.

**Conclusion:** For assessment of coronary stenosis during invasive coronary angiography a deep-learning-based model showed promising results for predicting visual assessment (ROC AUC of 0.903). Compared to previous work, our approach demonstrates performance increase, includes all 16 segments, does not exclude revascularized patients, is externally tested, and is simpler using fewer steps and fewer models.

## Introduction

X-ray multi-frame images (also known as cine loops, videos, views, or projections) acquired during an invasive coronary angiography (CAG) yield detailed information about the anatomy and flow in the coronary arteries.^1^ Cine loops are acquired separately for the left coronary artery (LCA) and right coronary artery (RCA), and views are acquired from different angulations. During and after recordings, coronary angiographies (CAGs) are visually assessed to identify and quantify stenosis on all 16 coronary artery segments^2^. This visual assessment, often called “eyeballing”, involves assessing the diameter reduction of the artery segment compared to the proximal reference in percentage. Based on the presence of stenoses, the need for pharmacotherapy and revascularization can be considered.^2^

The visual assessment of a stenosis has a high observer variance^2,3^. Recent guidelines suggest unnecessary use of percutaneous coronary intervention (PCI) and coronary artery bypass grafting (CABG) in 1-2% and 10-15% of cases, respectively, which is likely caused by inaccurate assessment of stenoses.^2–3^

Objective stenosis assessment can be evaluated by fractional flow reserve (FFR) measurements during the procedure, measuring the pressure drop across a stenosis to determine the hemodynamic significance of a stenosis. Despite the proven benefits of wire-based FFR measurements^4–9^, utilization varies across hospitals and countries, with a utilization span between 5-17%.^10–13^

Alternatively, quantitative coronary angiography (QCA) can be used for objective measurements of a vessel diameter reduction using image analysis software. QCA relies typically on keyframe extraction, manual segmentation of vessels with stenosis, followed by 3D reconstruction using two different angulations.^14–15^ While FFR is considered the ground truth for determining hemodynamically significant stenosis, QCA is attractive for research as it can be performed offline and after the CAG.^14^ It has been shown that revascularization of non-culprit lesions based on QCA can reduce future incidences of myocardial infarction.^15^ Unfortunately, both QCA and FFR are expensive, time-consuming, and require special training to produce reliable results.

Considering these challenges, there has been growing interest in applying deep-learning-based methodologies for automatic stenosis estimation.^16–23^ Previous, similar work involving reasonably sized datasets presents a complex pipeline having six steps and eight models, focusing only on 11 segments, and excludes patients with prior revascularization. ^22–23^

In this paper, we present an end-to-end learning-based approach aiming to provide a useful clinical tool. Our method has improved performance compared to related work, capable of estimating stenosis on all 16 segments without exclusion of patients with prior revascularization, and the performance was evaluated on an external test set from a different hospital. Furthermore, the performance was evaluated against both visual assessments, QCA, and FFR.

## Methods

### Cohort description

#### Rigshospitalet dataset: Cohort description

Our dataset used for model development and testing included 19,414 patients, comprising 332,582 X-ray cine loops, were extracted from Rigshospitalet, Copenhagen, (period 2006- 2016). In total, the dataset contained 23,415 CAGs, and each CAG contained an average of 17.8 cine loops. The characteristics of the 19,414 patients, corresponding to the time point of coronary angiography, are presented in Table 1. CAGs were recorded using Philips Medical Systems, GE HealthCare, and Siemens Healthineers angio systems. The CAGs were linked to the Eastern Denmark Heart Registry (EDHR) database. The EDHR database contains information about visual assessment in each of the three major coronary arteries, reported according to the 16-segment classification protocol.^25–27^ Additionally, the indication for coronary angiography and the treatment was recorded (Supplemental Table S1). Segments displaying borderline or intermediate stenosis were, if appropriate, further evaluated using Fractional Flow Reserve (FFR). Every entry in the EDHR database was manually registered by interventional cardiologists as part of clinical practice. We used 14,358 randomly selected patients for model development (90% for the training set with 12,846 patients and 10% for the validation set with 1,389 patients). For evaluation of the model performance, we used 5,056 randomly selected patients for evaluating the performance, which we will refer to as the internal test set (Supplemental Table S2).

**Table 1.** Cohort characteristics.

| Features | Total |
| --- | --- |
| Patients | 19,414 |
| Age, years | 67.3 ± 12.4 |
| Males (%) | 13,377 (68.9%) |
| Diabetes (%) | 3,634 (18.7%) |
| Hypertension (%) | 10,042 (51.7%) |
| Smokers and ex-smokers (%) | 12,752 (65.7%) |
| No vessel abnormalities (%) | 2,892 (14.9%) |
| Atheromatous vessels (%) | 4,886 (25.2%) |
| 1 vessel disease (%) | 7,030 (36.2%) |
| 2 vessel disease (%) | 3,477 (17.9%) |
| 3 vessel disease (%) | 3,209 (16.5%) |
| Left main disease (%) | 1094 (5.64%) |
| Prior PCI (%) | 4,124 (21.2%) |
| Prior CABG (%) | 1,506 (7.7%) |
| Right dominant (%) | 15,795 (81.4%) |
| Left dominant (%) | 1,811 (9.3%) |
| Co dominant (%) | 2,023 (10.4%) |
| Arrhythmia device (%) | 827 (4.3%) |

#### Skejby Hospital: Cohort for external testing

We further evaluated the model on 608 patients from Skejby Hospital in the Central Denmark Region, which we refer to as the external test set. These patients were selected following initial findings of suspicious stenosis from coronary computed tomography angiography (CTA). Each patient had a single coronary angiography recorded using Philips Medical Systems and Siemens Healthineers Angio System scanners. FFR was measured in all segments technically feasible for FFR measurements. All applicable segments were also analyzed with QCA using Medis QAngio®XA 3D, Netherlands.

### Overview: A deep learning-based approach for automated stenosis estimation

We employed a multi-stage approach for estimating the degree of stenosis on all coronary artery segments. First, we manually annotated a subset of cine loops as either left or right coronary arteries (LCA/RCA). Secondly, we developed and trained a deep learning model to differentiate between LCA and RCA in coronary angiography cine loops. This model was used to classify all cine loops as LCA, RCA, or “other”. Thirdly, we selected all cine loops before revascularization using an automated approach based on the classified cine loops and the timestamp. Fourthly, we developed two deep learning models for estimating stenosis: one for LCA and another for RCA, utilizing the cine loops before revascularization. An overview of the approach can be found in the Central Illustration.

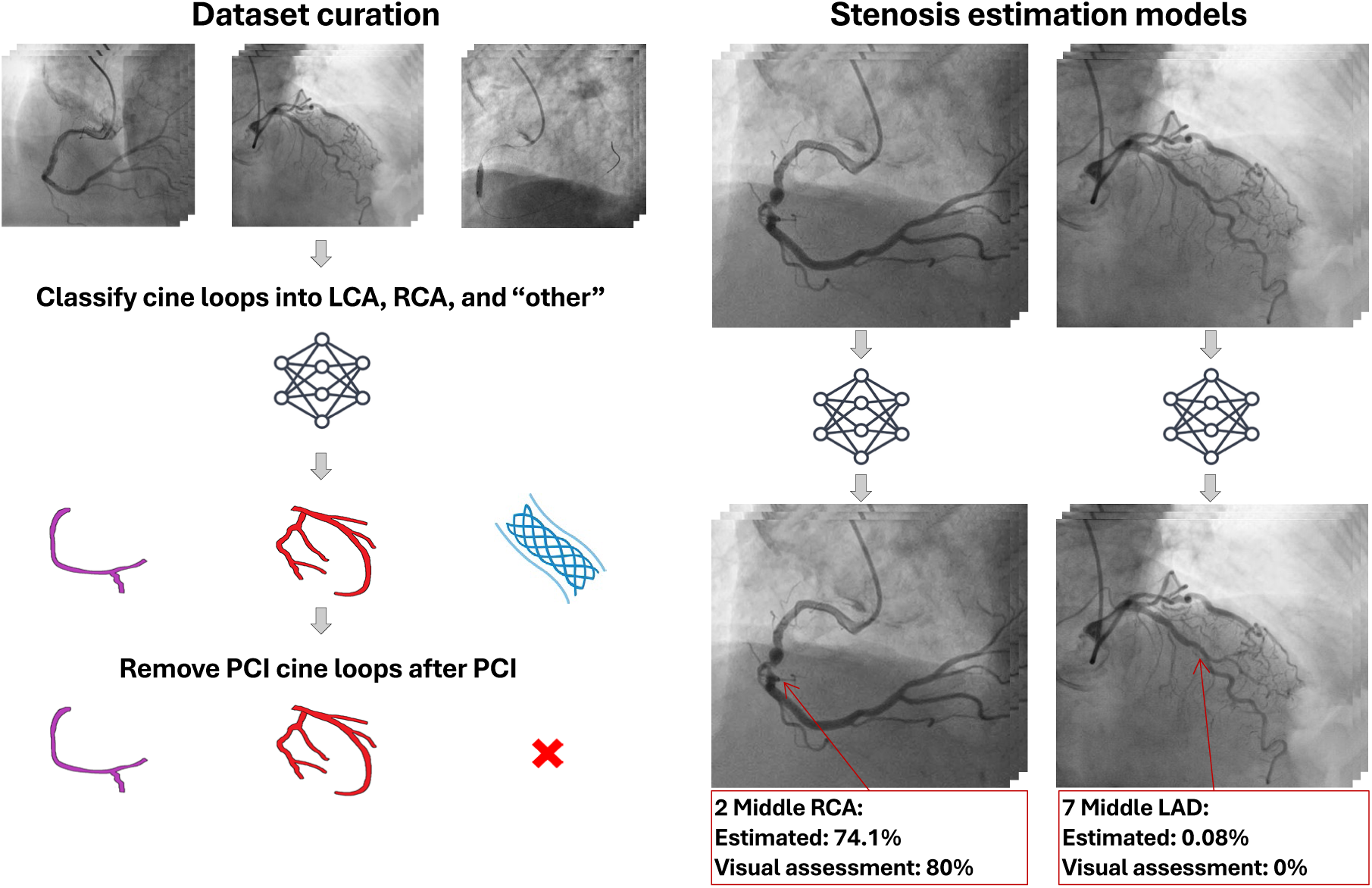
Central Illustration. Overview of the proposed approach for stenosis estimation.

### R2D+1 backbone deep learning model

We used the R2D+1 deep learning model,^29^ a supervised 3D convolutional neural network (CNN), which has previously been demonstrated state-of-the-art performance for CAG cine loops classification of right and left coronary artery.^28^ The R2D+1 model uses the R2D+1 block, which compresses the 3D convolutional block into a spatial block with filters of size 3 × 3 × 1 and a temporal block with filters of size 1 × 1 × 3. Non-linear ReLu activation is used between the spatial and temporal filters. The R2D+1 block can be interpreted as a combination of spatial and temporal filters, but with non-linearity between the two operations, extracting non-linear relations between spatial and temporal features.

For both CAG cine loop classification tasks and stenosis estimation, we employed the R2D+1 network. The model takes a CAG cine loop as input from the training set and learns discriminative features. The discriminatory features will depend on the target used for the model, and thus, the model learns different features for the cine loop classification model and stenosis estimation models.

### Annotation of cine loops

To categorize all the cine loops, we manually labelled a subset of 18,058 cine loops from 1,228 patients as LCA, RCA and “other”. The “other” category included cine loops, in which the LCA or the RCA was not present. The purpose of the “other” category was to exclude cine loops not relevant for visual assessment. We specifically categorized cine loops containing guide wires as “other”, even when they also displayed either the left or the right coronary artery. Cine loops containing chronic total occlusions (CTO) were still annotated as LCA or RCA. For training and validation, we used 1,047 patients with 15,068 cine loops. For model evaluation, we used a test dataset of 2,990 cine loops from 179 patients (Supplemental Table S2).

### Cine loop classification model

We developed a deep learning classification model designed to classify cine loops into one of the three categories: LCA, RCA and “other” using the labeled subset. We used the trained cine loop classification model to categorize the cine loops in the training/validation and the test sets as LCA (LAD and LCX), RCA and “other”. This classification step extends the work of Eschen et al.^28^, who focused on left and right coronary artery classification, by incorporating an additional “other” category.

### Diagnostic cine loop selection

The cine loops obtained during, and post revascularization are not applicable to the decision-making process regarding revascularization in a deployment scenario of the models. Additionally, cine loops obtained during, and post revascularization are highly associated with stenoses and may, therefore, introduce bias in the model during training. Consequently, we excluded cine loops performed during and post revascularization procedures. This exclusion involved removing cine loops categorized as “other” and any cine loops obtained after this category appeared in the sequence. We denote this step as the “diagnostic cine loop selection step” as depicted in the Central Illustration (see also Supplemental Methods 1). A detailed explanation of the data inclusion process is presented in Supplemental Materials Section 1.1, and Figure S1.

### Training the stenosis estimation models

Using the diagnostic cine loop selection procedure, we included cine loops of LCA and RCA and excluded cine loops obtained during and after PCI. The selection procedure resulted in 13,284 patients with 31,161 RCA cine loops, and 13,768 patients with 62,165 LCA cine loops (see Supplemental Materials Table S3, and Figure S2-S3 for details).

We developed the stenosis estimation models individually for RCA and LCA using 31,161 and 62,165 cine loops. For both models, we used multi-output regression models. For the RCA stenosis estimation model, the final linear layer contained five neurons, one for each of the five RCA segments. Specifically, for the RCA model, the five output neurons corresponded to artery segments relevant to the RCA. Similarly, for the LCA stenosis estimation model, we used a multi-output regression model with 13 neurons in the final linear layer, one for each of the 13 segments relevant to the LCA (we also include the Posterior Descending Artery (PDA) and the Posterior Left Ventricular Artery (PLA) in the LCA model). This design ensures that the model can simultaneously make stenosis estimates for each segment, making it capable of handling multiple stenoses at once.

As the visual stenosis assessment was only reported for segments with potentially significant stenoses, we replaced the missing values with zeros as these were missing by purpose. Therefore, we had a complete dataset that included cine loops and corresponding visual assessment of stenosis on all coronary artery segments.

### Evaluating stenosis estimation models against visual assessment

Using the diagnostic cine loop selection procedure, we established a test set with 5,056 patients (24,359 cine loops of the LCA from 5,015 patients and 12,138 cine loops of the RCA from 4,788 patients, as shown in Supplemental Figure S4). Additionally, we leveraged the external cohort with 608 patients for external validation (2,949 cine loops of LCA from 608 patients and 1,425 cine loops of RCA from 599 patients as depicted in Supplemental Figure S5).

The final LCA and RCA stenosis estimates were obtained by selecting the most severe stenosis estimate (the maximum stenosis) from all cine loops in a CAG examination. Coronary dominance was used to decide whether LCA or RCA predictions should be employed to evaluate the PDA and PLA segments.

We evaluated the model’s ability to predict diameter stenosis as a continuous outcome. We also assessed its ability to distinguish between significant and non-significant stenosis as a binary outcome. We applied the clinical threshold for significant coronary artery diameter stenosis >70%, except for the left main segment, which was >50%.^27^ We assessed the performance of the stenosis predictions for each of the 16 segments of the LCA and RCA models, as well as the overall average performance.

We also evaluated the stenosis estimation model using our “Angin-FFR Subset”. The “Angina FFR Subset” was part of the internal test set, but consists of patients with similar characteristics as the patients in the external test set. Hence, this subset included 499 patients with indications of ischemia and angina, patients with FFR measurements in at least one segment, patients with atheromatous lesions, and those with single-vessel and two-vessel disease.

### Evaluating stenosis estimation models against FFR

For the subset of angiographies followed by FFR measurements (1180 patients in the internal test set and 439 patients in the external test set), we compared the stenosis estimates against FFR measurements. The FFR measurements were transformed to a comparable scale by subtracting the FFR measurements from one. We evaluated the performance on detecting hemodynamic significant stenosis (FFR ≤ 0.8). To establish a comparable baseline for predicting FFR ≤ 0.8, we evaluated the performance using visual assessments as predictors.

### Evaluating stenosis estimation models against QCA

The estimated stenosis was also compared against QCA in the external test set for 359 patients. The evaluation was performed similarly to the evaluation against visual assessment. As we had access to both the visual assessments and FFR in this dataset for 209 of the spatients, we established a baseline for comparison using visual assessment and FFR as predictors for QCA.

### Statistical analysis

The estimated stenosis was compared against visual assessment, FFR, and QCA measurements using mean absolute error (MAE) and Pearson’s correlation coefficient (r). The estimated stenoses were also compared against FFR using these metrics.

To evaluate the performance on detecting significant stenoses, we used the area under the Receiver Operating Characteristic curve (ROC AUC), the area under the precision-recall curve (PR AUC), F1 score, precision, sensitivity, and specificity. The confidence intervals were computed using 1000 bootstrap samples at a 95% confidence level.

### Approvals and data availability

Approval for data access was granted by the National Committee on Health Research Ethics (1708829 “Genetics of cardiovascular disease”, ID P-2019-93), The Danish Data Protection Agency (ref: 514-0255/18-3000, 514-0254/18-3000, SUND-2016-50), and by the Danish Patient Safety Authority (3-3013-1731-1, appendix 31-1522-23). All personal identifiers were pseudo-anonymized. Data access applications can be made to the Danish Health Data Authority (contact). Anyone wanting access to the data and to use them for research will be required to meet research credentialing requirements as outlined at the authority’s web site: https://sundhedsdatastyrelsen.dk/da/english/health_data_and_registers/research_services

. Requests are normally processed within 3 to 6 months.

The source code for this study is available (URL to come).

## Results

### Performance of the cine loop classification model

The performance of the cine loop classification model had a macro F1 score of 0.972 (95% CI: 0.972-0.972) on the internal test set (Figure S5 in Supplemental Materials). We assessed the discordant predictions (79 cine loops) and found that most of these originated from cases with ambiguous labels (e.g., cine loops obtained while measuring the FFR using a guide wire).

### Performance of the stenosis estimation model

For predicting the visual assessment (diameter stenosis), we obtained a MAE of 0.178 (95% CI 0.177-0.179), and a Pearson’s correlation coefficient of 0.661 (95% CI 0.656-0.666) on the internal test set. On the “Angina-FFR Subset” we obtained an MAE of 0.156 (95% CI: 0.144- 0.168), Pearsons’s correlation coefficient of 0.293 (95% CI: 0.196-0.393) when predicting visual assessment. On the external test set, we obtained an MAE of 0.186 (95% CI: 0.182- 0.190) and a Pearson’s correlation coefficient of 0.386 (95% CI: 0.317-0.373) compared to the visual assessment.

We evaluated the model’s performance on significant stenosis identification and obtained a ROC AUC of 0.903 (95% CI: 0.900-0.906), and PR AUC of 0.693 (95% CI: 0.685-0.701), as seen in Figure 1. On the “Angina-FFR Subset” we obtained a ROC AUC of 0.849 (95% CI: 0.829- 0.867), PR AUC of 0.486 (95% CI: 0.436-0.530) when predicting significant stenoses.

**Figure 1.**
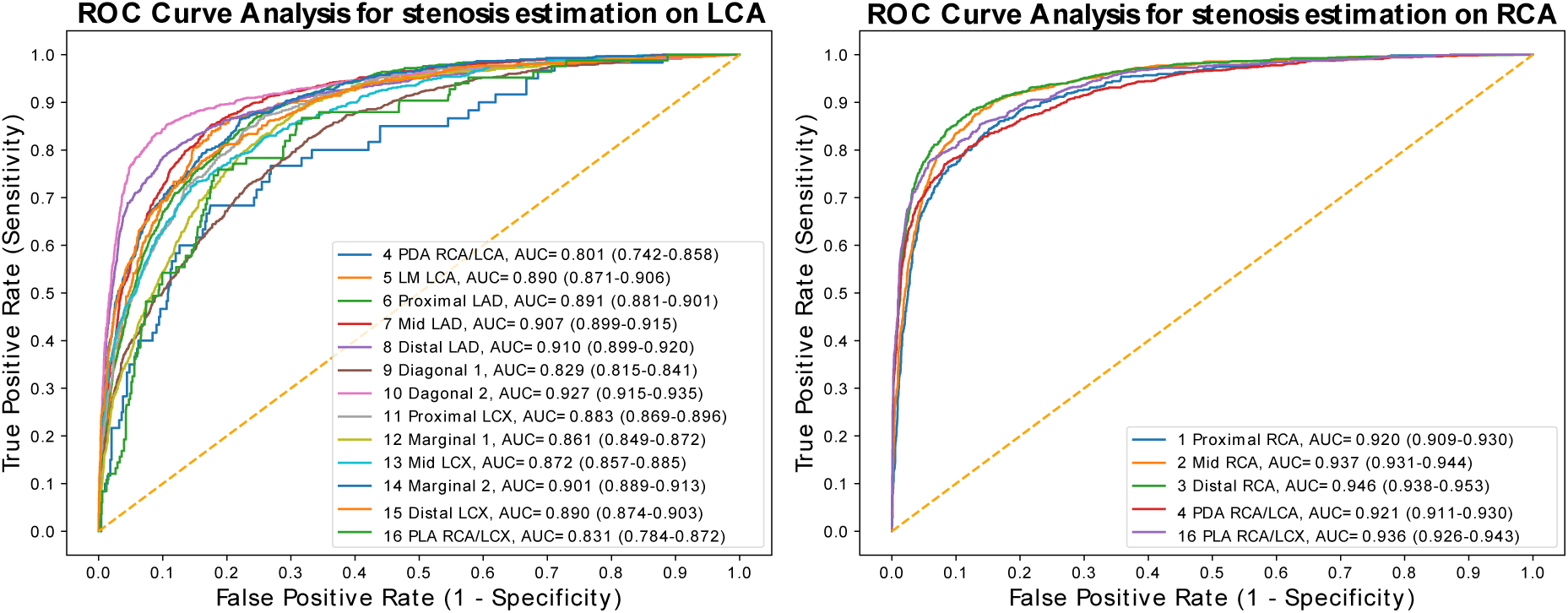
ROC curve for significant stenosis detection for each segment on the internal test set (visual assessment of diameter stenosis >70%).

For detection of significant stenosis on the external test set, the ROC AUC decreased to 0.833 (95% CI: 0.814-0.852), and PR AUC decreased to 0.219 (95% CI: 0.190-0.250) as shown in Table 2 (the performances on the individual segments are depicted in Supplemental Materials Table S5-S6).

**Table 2.** Performance on predicting visual assessment.

|  | Internal test set | External test set |
| --- | --- | --- |
| Method | Estimated stenosis (ours) | Estimated stenosis (ours) |
| MAE | 0.178 (0.177-0.179) | 0.186 (0.182-0.190) |
| r | 0.661 (0.656-0.666) | 0.345 (0.317-0.373) |
| ROC AUC | 0.903 (0.900-0.906) | 0.833 (0.814-0.852) |
| PR AUC | 0.693 (0.685-0.701) | 0.219 (0.190-0.250) |
| F1 | 0.637 (0.631-0.643) | 0.314 (0.284-0.343) |
| Sensitivity | 0.681 (0.674-0.689) | 0.548 (0.503-0.593) |
| Specificity | 0.922 (0.920-0.924) | 0.901 (0.895-0.907) |
| Precision | 0.599 (0.591-0.606) | 0.220 (0.199 -0.245) |

### Predicting fractional flow reserve

For predicting the measured FFR values on the internal test set, we obtained an MAE of 0.157 (95% CI 0.148-0.165) and a Pearson’s correlation coefficient of 0.220 (95%CI 0.163- 0.281). On the external test set, we obtained an MAE of 0.120 (0.111-0.129) and a Pearson’s correlation coefficient of 0.441 (95% CI 0.375-0.502) when predicting the measured FFR values.

For the detection of hemodynamically significant stenosis (FFR≤0.80), we obtained a ROC AUC of 0.651 (95% CI: 0.616-0.686) on the internal test set, and a ROC AUC of 0.780 (95% CI: 0.743-0.817) on the external test set as shown in Table 3 (performance on individual segments are depicted in Tables S7-S10 in Supplemental Materials.

**Table 3.** Performance on predicting FFR.

|  | Internal test set |  | External test set |  |  |
| --- | --- | --- | --- | --- | --- |
| Method | Estimated stenosis<br>(this paper) | Visual<br>assessment | Estimated stenosis<br>(this paper) | Visual<br>assessment | QCA |
| MAE | 0.157 (0.148-0.165) | 0.394 (0.385-0.401) | 0.120 (0.111-0.129) | 0.254 (0.240-0.268) | 0.265 (0.248-0.285) |
| r | 0.220 (0.163-0.281) | 0.549 (0.494-0.598) | 0.441 (0.375-0.502) | 0.640 (0.600-0.676) | 0.180 (0.037-0.325) |
| ROC AUC | 0.651 (0.616-0.686) | 0.853 (0.828-0.876) | 0.780 (0.743-0.817) | 0.844 (0.817-0.877) | 0.575 (0.491-0.663) |
| PR AUC | 0.400 (0.353-0.452) | 0.635 (0.583-0.685) | 0.441 (0.365-0.524) | 0.532 (0.449-0.606) | 0.430 (0.318-0.552) |
| F1 | 0.417 (0.368-0.465) | 0.680 (0.636-0.723) | 0.438 (0.361-0.511) | 0.558 (0.497-0.621) | 0.058 (0.000-0.143) |
| Sensitivity | 0.411 (0.360-0.465) | 0.690 (0.638-0.742) | 0.386 (0.311-0.464) | 0.611 (0.526-0.689) | 0.030 (0.000-0.077) |
| Specificity | 0.786 (0.761-0.814) | 0.869 (0.847-0.892) | 0.914 (0.891-0.936) | 0.868 (0.842-0.894) | 0.985 (0.962-1.000) |
| Precision | 0.427 (0.372-0.480) | 0.670 (0.619-0.716)) | 0.510 (0.423-0.594) | 0.519 (0.445-0.590) | 0.488 (0.000-1.000) |

### Predicting QCA

We further evaluated the performance on QCA prediction on the external test set (QCA was not measured in the dataset from Rigshospitalet). We obtained a MAE of 0.210 (95% CI 0.203-0.217) and a Pearson’s correlation coefficient of 0.477 (95% CI 0.423-0.530). On detection QCA diameter stenosis >70%, we obtained a ROC AUC of 0.798 (95% CI: 0.782- 0.814), as depicted in Table 4. On detecting QCA-based significant stenosis, our models were consistently better than visual assessment and FFR with a ROC AUC of 0.798 versus 0.658 and 0.575 (additional performance metrics on individual segments are depicted in Table S11-S12 in Supplemental Materials).

**Table 4.** Performance for predicting QCA on the external test set.

| Method | Estimated stenosis (this paper) | Visual assessment | FFR |
| --- | --- | --- | --- |
| MAE | 0.210 (0.203-0.217) | 0.351 (0.343-0.360) | 0.265 (0.248-0.285) |
| r | 0.477 (0.423-0.530) | 0.358 (0.302-0.408) | 0.180 (0.037-0.325) |
| ROC AUC | 0.798 (0.741-0.842) | 0.658 (0.591-0.726) | 0.575 (0.491-0.663) |
| PR AUC | 0.340 (0.243-0.443) | 0.273 (0.179-0.374) | 0.430 (0.318-0.552) |
| F1 | 0.246 (0.188-0.304) | 0.246 (0.183-0.312) | 0.058 (0.000-0.143) |
| Sensitivity | 0.578 (0.471-0.688) | 0.446 (0.340-0.563) | 0.030 (0.000-0.077) |
| Specificity | 0.817 (0.797-0.836) | 0.872 (0.853-0.889) | 0.985 (0.962-1.000) |
| Precision | 0.157 (0.116-0.198) | 0.170 (0.123-0.225) | 0.488 (0.000-1.000) |

## Discussion

Our deep learning model demonstrated robust performance in classifying cine loops into LCA, RCA, and “other” categories, with a macro F1 score of 0.972. For detecting significant stenosis, high ROC AUC levels of 0.903 on the internal test set and of 0.833 on the external test set were found. The model outperformed visual assessment when validated against QCA, achieving a ROC AUC of 0.798. For predicting hemodynamically significant stenosis measured by FFR, the model achieved a ROC AUC of 0.651 on the internal test set and 0.780 on the external test set. Here, we discuss our findings regarding the related works, visual assessments, FFR, and QCA, and finally, we address the limitations of our approach.

### Related works

In recent years, several studies have focused on the significant stenosis detection in coronary angiography (CAG) cine loops. As mentioned, although many of these advancements have been based on small datasets only considering single CAG frames^16–18^, efforts for significant stenosis detection on larger datasets exist^19–23^. For instance, Avram et al. curated and trained a model (CathAI) including 11,972 patients, achieving a ROC AUC of 0.839 on an internal test set.^19^ Most recently, and comparable to our work, Langlais et al. introduced DeepCoro, developed by the same research group behind CathAI. DeepCoro is a 6-step pipeline that includes primary structure identification, stenosis detection, frame registration, coronary artery segmentation, alignment of stenosis with segments, and finally, stenosis regression.^23^ DeepCoro was developed using 182,418 coronary angiography cine loops, and it obtained a ROC AUC of 0.829 on stenosis detection, and a MAE of 20.15% on predicting visual assessment in percentage on an internal test set.

Compared to our results, DeepCoro achieved a test ROC AUC of 0.8294 (0.8215–0.8373) and a PR AUC of 0.5239 (0.5041–0.5421), which is significantly lower than our test performances of ROC AUC of 0.903 (0.900-0.906) and a PR AUC of 0.693 (0.685-0.685). Notably, our approach involves 2 steps and 3 models, while DeepCoro uses 6 steps and 8 models.^23^

Notably, the most similar work, i.e., DeepCoro only focused on 11 Segments, instead of 16 segments and excluded patients with prior CABG and PCI.^23^ Both aspects are highly relevant for assessing a CAG.^2^ Moreover, previous work did not evaluate the performance on both QCA, FFR, nor was the performance assessed on an external test set from another cohort and hospital. Finally, our methods can run on the fly with a processing speed of 0.03 seconds for a cine loop which is significantly better than DeepCoro with a processing speed of 62.6 seconds.

Hence, we aimed to address the limitations in existing work, and our models obtain superior performance, and the approach uses a simpler and faster pipeline.

### Comparison against visual assessment

While we obtained the best performance reported in the literature for significant stenosis detection (ROC AUC of 0.903 and PR AUC of 0.693), we observed a notable performance drop on the external test set (ROC AUC of 0.833 and PR AUC of 0.219). A similar pattern emerged when evaluating the model on the “Angina-FFR Subset,” where the ROC AUC decreased to 0.849 and the PR AUC to 0.486. The most significant factor contributing to this performance decline appears to be the difference in patient characteristics. The external test set consisted of patients selected based on prescreening with CTA, leading to a higher proportion of individuals with intermediate stenosis and excluding those with mild stenosis or multivessel disease, such as patients with STEMI or NSTEMI.

### Comparison against FFR

For predicting hemodynamically significant stenoses, our model achieved a ROC AUC of 0.651 (95% CI: 0.616-0.686) on the internal test set and 0.780 (95% CI: 0.743-0.817) on the external test set. The performance of the model was inferior on predicting hemodynamical significant stenosis using visual assessment achieving a ROC AUC of 0.853 and 0.844 on the test set and the external test set. However, the reported visual assessments can be overoptimistic and biased towards FFR measurements, as they are typically reported after the FFR is measured and are not blinded to the FFR measurement. Another notable finding is that using QCA to determine hemodynamically significant stenosis from FFR yielded low performance. While our models demonstrated good performance, there is still room for improvement for predicting hemodynamically significant stenosis.

### Comparison against QCA

For detecting the clinically important threshold of QCA diameter stenosis >70% on the external test set, our model achieved a ROC AUC of 0.798 (95% CI: 0.782-0.814). This performance was consistently better than visual assessment, which obtained a ROC AUC of 0.658 (95% CI: 0.591-0.726), and FFR, which obtained a ROC AUC of 0.575 (95% CI: 0.491-0.552). While there is strong evidence that FFR is optimal for revascularization decisions, QCA is attractive for research. Our methodology has the potential to be used as a fast and accurate alternative to traditional QCA.

### Limitations

Despite the promising results, we acknowledge some limitations in our results. The most notable limitation is that the stenosis estimation models were trained on patients undergoing routine coronary angiography, including patients without disease and those with multivessel disease. As a result, the stenosis estimation models are not guaranteed to generalize to patient cohorts with other inclusion/exclusion criteria (e.g., patients undergoing CTA before coronary angiography), which can be seen in the decreased performance on predicting visual assessment in the external test set and the subset “Angina-FFR Subset”. Secondly, the stenosis estimation models were trained using visual assessments, as we did not have access to QCA, and FFR measurements which were only available for borderline stenosis segments and where it was technically feasible to perform the measurements.

## Conclusion

Our approach for stenosis estimation showed promising results, outperforming previous work on predicting visual assessments. However, a significant performance drop was observed in the external test cohort, which had suspected stenosis detected by CTA. Predicting hemodynamically significant stenosis measured by FFR using the stenosis estimation models did not surpass using visual assessments as predictors, indicating that improvements in this area are likely needed. Notably, the stenosis estimations were better at predicting QCA diameter stenosis compared to visual assessments. These results suggest that our approach for stenosis estimation is clinically relevant, offering a faster and more objective alternative to traditional methods. Future research should focus on improving the models and investigating the effect of the estimated values on the treatment compared with traditional methods.

## Clinical perspectives

A deep learning-based approach can estimate the degree of stenosis directly using cine loops. The model is the first of its kind to predict stenosis in all 16 coronary artery segments. The deep learning model demonstrated strong performance in predicting visual assessment of stenosis, and the model was better than traditional visual assessment in predicting stenosis measured. The model offers a fast and accurate alternative to QCA. However, further improvements are necessary to enhance its ability to determine hemodynamical significant stenosis.

## Data Availability

Data access applications can be made to the Danish Health Data Authority (contact). Anyone wanting access to the data and to use them for research will be required to meet research credentialing requirements as outlined at the authority's web site: https://sundhedsdatastyrelsen.dk/da/english/health_data_and_registers/research_services. Requests are normally processed within 3 to 6 months.

## Acknowledgements

This work was funded by the Novo Nordisk Foundation (NNF17OC0027594 and NNF14CC0001) and the Innovation Fund Denmark (518-00102B).

