## Supplemental materials for "Automated stenosis estimation of coronary angiographies using end-to-end learning"

### 1. Supplemental Methods

#### Training details

The R2D+1 models were trained using eight A100 GPUs with 40GB of memory, using the SmartCache protocol to maximize GPU utilization.^30^

For the cine loop classification model, an iteration was defined as a training step with 1,024 videos, with each GPU handling 128 videos. For the stenosis estimation models, an iteration was defined as a training step with 128 videos, with each GPU handling 16 videos. The smaller number of cached videos for stenosis estimation was due to more cached data in the validation set for this model. A replace rate of 20% was used to replace the cached data in each iteration.

The preprocessing steps involved resizing the videos to 224 × 224 pixels in the spatial domain and random cropping of video clips with a length of 32. Data augmentation) was performed online, including random rotation, random temporal scaling, random spatial scaling, and random translation.

The cine loop classification model was trained using a cross-entropy loss with a batch size of 8 pr. GPU for 5,000 iterations with an initial learning rate of 0.02. At 2,500 iterations, the learning rate decreased by a factor of 10.

The stenosis estimation models were trained similarly, yet we trained these models using iterations with the weighted focal L1 loss.^31^ The loss was weighted with the inverse frequency of the labels using label distribution smoothing tailored regression models.^32^ The cine loop classification model was trained for approximately 12 hours. The stenosis estimation models were trained for 32,000 iterations, and the learning rate was reduced by a factor of 10 after 16,000 iterations. The training time for the stenosis estimation models was approximately 24 hours. Throughout the training process, we monitored the performance on the validation set for each training iteration. Model selection was carried out by evaluating the loss on the validation set and selecting the model with the lowest loss. The weights were initialized with pre-trained weights from the Kinetics dataset.^33^

### 2. Supplemental Tables

#### 1.1 Table S1: Characteristics of the CAGs in the dataset from Rigshospitalet

|  | Total |
| --- | --- |
| CAGs | 23,415 |
| Patients | 19,414 |
| Cine loops | 332,582 |
| CAG indication: Angina (%) | 4,824 (20.6%) |
| CAG indication: STEMI (%) | 7,120 (30.4%) |
| CAG indication: NSTEMI (%) | 5,947 (25.4%) |
| CAG indication: Follow up PCI/CABG (%) | 1,372 (5.9%) |
| CAG indication: Other (%) | 4,152 (17.7%) |
| Treatment: No treatment (%) | 2,119 (9.0%) |
| Treatment: Pharmacotherapy (%) | 7.275 (31.1%) |
| Treatment: PCI (%) | 10315 (44.1%) |
| Treatment: CABG (%) | 1684 (7.2%) |
| Treatment: Missing information (%) | 850 (3.6%) |
| Treatment: Missing information on (%) | 128 (10.9%) |

#### 1.2 Table S2. Overview of the different subsets used for model development and testing

|  | Training set | Validation set | Internal test set | External test dataset |
| --- | --- | --- | --- | --- |
| Source | Rigshospitalet in Copenhagen, Denmark | | | Skejby Hospital in Aarhus, Central Denmark Region |
| Purpose | The model learns from the training data | Used for experiment with different hyperparameters.  Model selection using lowest loss (early stopping). | Used only for report performance.  Separated from training and at the beginning of the study | Used only for report performance on a external cohort (external validation). |
| Selection | Random selected (90%) from the union of training and validation data. | Random selected (10%) from the union of training and validation data. | Randomly selected | All samples are selected |
| Patients | 12,506 | 1,334 | 5,056 | 608 |
| Cine loops | 84,241 | 9,085 | 36,497 | 4,374 |

#### 1.3 Table S3: Dataset from Rigshospitalet used for developing cine loop classification model

|  | Cine loop classification model | | |
| --- | --- | --- | --- |
|  | Training | Validation | Test |
| CAGs | 1,173 | 133 | 232 |
| Patients | 944 | 105 | 179 |
| Cine loops | 13,511 | 1,557 | 2,990 |
| CAG indication: Angina (%) | 351 (29.9%) | 46 (34.6%) | 47 (20.3%) |
| CAG indication: STEMI (%) | 128 (10.9%) | 13 (9.8%) | 65 (28.0%) |
| CAG indication: NSTEMI/Unstable angina (%) | 336 (28.6%) | 35 (26.3%) | 71 (30.6%) |
| CAG indication: Follow up PCI/CABG (%) | 107 (9.1%) | 9 (6.8%) | 7 (3.0%) |
| CAG indication: Other (%) | 251 (21.4%) | 30 (22.6%) | 42 (18.1%) |
| Treatment: No treatment (%) | 142 (12.1%) | 15 (11.3%) | 24 (10.3%) |
| Treatment: Pharmacotherapy (%) | 561 (47.8%) | 66 (49.6%) | 69 (29.7%) |
| Treatment: PCI (%) | 292 (24.9%) | 27 (20.3%) | 94 (40.5%) |
| Treatment: CABG (%) | 50 (4.3%) | 6 (4.5%) | 23 (9.9%) |
| Treatment: Missing information on (%) | 128 (10.9%) | 14 (14.3%) | 22 (9.6%) |

#### 1.4 Table S4: Dataset used for developing stenosis estimation models

|  | Stenosis estimation models | | |
| --- | --- | --- | --- |
|  | Training | Validation | Test |
| CAGs | 14,747 | 1,570 | 6,342 |
| Patients | 12,506 | 1,334 | 5,056 |
| Cine loops | 84,241 | 9,085 | 36,497 |
| CAG indication: Angina (%) | 2912 (19.7%) | 334 (21.3%) | 1578 (24.9%) |
| CAG indication: STEMI (%) | 5236 (35.5%) | 541 (34.5%) | 1343 (21.2%) |
| CAG indication: NSTEMI/Unstable angina (%) | 3636 (24.7%) | 363 (23.1%) | 1948 (30.7%) |
| CAG indication: Follow up PCI/CABG (%) | 842 (5.7%) | 122 (7.8%) | 408 (4.4%) |
| CAG indication: Other (%) | 2121 (14.4%) | 220 (13.3%) | 1578 (24.9%) |
| Treatment: No treatment (%) | 1403 (9.5%) | 130 (8.3%) | 566 (8.9%) |
| Treatment: Pharmacotherapy (%) | 4223 (28.6%) | 473 (30.1%) | 2395 (37.8%) |
| Treatment: PCI (%) | 6979 (47.3%) | 744 (47.4%) | 2429 (38.3%) |
| Treatment: CABG (%) | 1025 (7.0%) | 115 (7.3%) | 406 (6.4%) |
| Treatment: Missing information on (%) | 1117 (7.6%) | 108 (6.9%) | 546 (8.6%) |

#### 1.5 Table S5. Performance on predicting visual assessment on internal test set using estimated stenosis

| Segments | CAGs | MAE | r | ROC AUC | PR AUC | F1 | Sensitivity | Specificity | Precision |
| --- | --- | --- | --- | --- | --- | --- | --- | --- | --- |
| 1 Proximal RCA | 5850 | 0.133 (0.129-0.138) | 0.651 (0.625-0.677) | 0.919 (0.908-0.930) | 0.652 (0.614-0.690) | 0.651 (0.621-0.682) | 0.637 (0.601-0.673) | 0.959 (0.954-0.964) | 0.668 (0.630-0.706) |
| 2 Middle RCA | 5850 | 0.171 (0.166-0.176) | 0.761 (0.744-0.776) | 0.937 (0.930-0.944) | 0.820 (0.796-0.841) | 0.755 (0.737-0.773) | 0.840 (0.820-0.861) | 0.893 (0.885-0.902) | 0.684 (0.663-0.706) |
| 3 Distale RCA | 5850 | 0.149 (0.145-0.154) | 0.772 (0.756-0.787) | 0.946 (0.937-0.952) | 0.845 (0.825-0.864) | 0.774 (0.755-0.793) | 0.805 (0.780-0.829) | 0.936 (0.929-0.943) | 0.746 (0.720-0.771) |
| 4 PDA | 5960 | 0.184 (0.179-0.189) | 0.711 (0.693-0.729) | 0.912 (0.903-0.921) | 0.789 (0.766-0.810) | 0.719 (0.697-0.739) | 0.690 (0.663-0.715) | 0.943 (0.937-0.950) | 0.751 (0.726-0.776) |
| 5 LM | 6244 | 0.094 (0.091-0.098) | 0.456 (0.425-0.486) | 0.890 (0.872-0.907) | 0.392 (0.343-0.447) | 0.006 (0.000-0.018) | 0.003 (0.000-0.009) | 1.000 (1.000-1.000) | 0.625 (0.000-1.000) |
| 6 Proximal LAD | 6244 | 0.238 (0.233-0.242) | 0.604 (0.584-0.624) | 0.891 (0.880-0.900) | 0.632 (0.601-0.661) | 0.579 (0.558-0.604) | 0.771 (0.745-0.796) | 0.832 (0.821-0.842) | 0.463 (0.439-0.486) |
| 7 Middle LAD | 6244 | 0.247 (0.243-0.252) | 0.684 (0.668-0.700) | 0.907 (0.899-0.915) | 0.760 (0.739-0.783) | 0.676 (0.659-0.693) | 0.881 (0.865-0.898) | 0.779 (0.767-0.791) | 0.547 (0.526-0.568) |
| 8 Distal LAD | 6244 | 0.181 (0.176-0.186) | 0.700 (0.680-0.719) | 0.910 (0.900-0.920) | 0.750 (0.722-0.777) | 0.704 (0.681-0.725) | 0.729 (0.702-0.755) | 0.931 (0.924-0.938) | 0.681 (0.655-0.708) |
| 9 Diagonal 1 | 6244 | 0.225 (0.220-0.230) | 0.512 (0.486-0.537) | 0.829 (0.815-0.843) | 0.531 (0.497-0.564) | 0.477 (0.450-0.505) | 0.412 (0.380-0.443) | 0.940 (0.933-0.946) | 0.567 (0.530-0.603) |
| 10 Diagonal 2 | 6244 | 0.153 (0.148-0.158) | 0.721 (0.702-0.741) | 0.927 (0.917-0.938) | 0.767 (0.739-0.794) | 0.730 (0.708-0.752) | 0.788 (0.762-0.812) | 0.936 (0.930-0.943) | 0.681 (0.653-0.708) |
| 11 Proximal LCX | 6244 | 0.175 (0.171-0.179) | 0.537 (0.511-0.562) | 0.883 (0.869-0.895) | 0.519 (0.480-0.557) | 0.489 (0.455-0.523) | 0.510 (0.471-0.551) | 0.940 (0.934-0.946) | 0.471 (0.433-0.509) |
| 12 Marginal 1 | 6244 | 0.227 (0.223-0.232) | 0.524 (0.500-0.547) | 0.861 (0.848-0.873) | 0.524 (0.489-0.558) | 0.508 (0.482-0.535) | 0.565 (0.531-0.596) | 0.892 (0.884-0.900) | 0.462 (0.434-0.491) |
| 13 Middle LCX | 6244 | 0.179 (0.175-0.183) | 0.561 (0.534-0.587) | 0.872 (0.858-0.885) | 0.553 (0.514-0.590) | 0.509 (0.480-0.543) | 0.547 (0.510-0.585) | 0.931 (0.925-0.937) | 0.478 (0.439-0.515) |
| 14 Marginal 2 | 6244 | 0.163 (0.159-0.168) | 0.582 (0.555-0.609) | 0.901 (0.888-0.913) | 0.587 (0.544-0.629) | 0.553 (0.520-0.586) | 0.617 (0.574-0.654) | 0.936 (0.929-0.942) | 0.500 (0.466-0.537) |
| 15 Distal LCX | 6244 | 0.153 (0.148-0.158) | 0.579 (0.550-0.607) | 0.890 (0.876-0.904) | 0.573 (0.528-0.616) | 0.543 (0.505-0.576) | 0.585 (0.545-0.626) | 0.943 (0.937-0.949) | 0.507 (0.468-0.547) |
| 16 PLA | 5985 | 0.166 (0.161-0.171) | 0.700 (0.679-0.719) | 0.916 (0.905-0.925) | 0.757 (0.730-0.783) | 0.704 (0.681-0.727) | 0.695 (0.666-0.723) | 0.943 (0.936-0.949) | 0.715 (0.688-0.742) |

#### 1.6 Table S6. Performance on predicting visual assessment on external test set using estimated stenosis

| Segments | CAGs | MAE | r | ROC AUC | PR AUC | F1 | Sensitivity | Specificity | Precision |
| --- | --- | --- | --- | --- | --- | --- | --- | --- | --- |
| 1 Proximal RCA | 593 | 0.149 (0.135-0.164) | 0.394 (0.258-0.520) | 0.816 (0.687-0.918) | 0.405(0.208-0.600) | 0.362 (0.233-0.487) | 0.706 (0.500-0.889) | 0.906 (0.884-0.936) | 0.245 (0.143-0.350) |
| 2 Middle RCA | 594 | 0.215 (0.197-0.233) | 0.529 (0.439-0.609) | 0.882 (0.831-0.927) | 0.464 (0.342-0.587) | 0.467 (0.383-0.544) | 0.848 (0.759-0.932) | 0.803 (0.768-0.838) | 0.322 (0.248-0.395) |
| 3 Distale RCA | 593 | 0.175 (0.157-0.194) | 0.358 (0.241-0.466) | 0.907 (0.820-0.963) | 0.268 (0.134-0.424) | 0.289 (0.180-0.406) | 0.895 (0.733-1.000) | 0.868 (0.839-898) | 0.174 (0.099-0.256) |
| 4 PDA | 595 | 0.190 (0.173-0.209) | 0.099 (0.019-0.192) | 0.757 (0.660-0.844) | 0.074 (0.037-0.126) | 0.086 (0.021-0.170) | 0.237 (0.050-0.467) | 0.878 (0.852-0.905) | 0.056 (0.012-0.114) |
| 5 LM | 599 | 0.102 (0.094-0.112) | 0.402 (0.279-0.515) | 0.858 (0.758-0.935) | 0.323 (0.173-0.493) |  |  |  |  |
| 6 Proximal LAD | 597 | 0.270 (0.258-0.283) | 0.460 (0.379-0.538) | 0.810 (0.761-0.858) | 0.521 (0.423-0.624) | 0.517 (0.444-0.595) | 0.656 (0.566-0.746) | 0.813 (0.780-0.848) | 0.426 (0.353-0.497) |
| 7 Middle LAD | 597 | 0.307 (0.293-0.322) | 0.319 (0.230-0.405) | 0.778 (0.722-0.830) | 0.385 (0.274-0.499) | 0.359 (0.291-0.432) | 0.721 (0.613-0.819) | 0.663 ( 0.625-0.703) | 0.239 (0.182-0.293) |
| 8 Distal LAD | 600 | 0.172 (0.160-0.185) | 0.173 (0.066-0.291) | 0.761 (0.652-0.853) | 0.112 (0.043-0.219) | 0.187 (0.062-0.320) | 0.351 (0.111-0.589) | 0.994 (0.913-0.951) | 0.134 (0.044-0.240) |
| 9 Diagonal 1 | 601 | 0.191 (0.182-0.201) | 0.060 (0.003-0.124) | 0.637 (0.514-0.739) | 0.051 (0.022-0.097) | 0.050 (0.000-0.164) | 0.057 (0.000-0.211) | 0.963 (0.948-0.978) | 0.045 (0.000-0.150) |
| 10 Diagonal 2 | 601 | 0.139 (0.127-0.152) | 0.000 (0.000-0.000) | 0.517 (0.034-0.779 | 0.012 (0.012-0.026) | 0 (0-0) | 0.000 (0.000-0.000) | 0.933 (0.913-0.951) | 0.000 (0.000-0.000) |
| 11 Proximal LCX | 600 | 0.184 (0.173-0.195) | 0.364 (0.224-0.485) | 0.815 (0.720-0.898) | 0.350 (0.176-0.518) | 0.363 (0.212-0.510) | 0.389 (0.212-0.577) | 0.964 ( 0.946-0.979) | 0.341 (0.179-0.515) |
| 12 Marginal 1 | 600 | 0.220 (0.209-0.231) | 0.307 (0.215-0.407) | 0.862 (0.791-0.920) | 0.215 (0.112-0.357) | 0.307 (0.179-0.427) | 0.541 (0.333-0.735) | 0.919 (0.896-0.941) | 0.215 (0.121-0.328) |
| 13 Middle LCX | 601 | 0.194 (0.182-0.207) | 0.247 (0.136-0.353) | 0.780 (0.684-0.861) | 0.206 (0.114-0.324) | 0.316 (0.182-0.442) | 0.352 (0.194-0.529) | 0.949 (0.930-0.966) | 0.294 (0.167-0.436) |
| 14 Marginal 2 | 601 | 0.161 (0.150-0.174) | 0.072 (0.025-0.125) | 0.785 (0.660-0.871) | 0.045 (0.018-0.083) | 0 (0-0) | 0.000 (0.000-0.000) | 0.939 (0.919-0.957) | 0.000 (0.000-0.000) |
| 15 Distal LCX | 601 | 0.149 (0.138-0.160) | 0.000 (0.000-0.000) | 0.790 (0.755-0.822) | 0.013 (0.007-0.031) | 0 (0-0) | 0.000 (0.000-0.000) | 0.942 (0.923-0.960) | 0.000 (0.000-0.000) |
| 16 PLA | 597 | 0.157 (0.140-0.174) | 0.123 (0.007-0.253) | 0.731 (0.573-0.861) | 0.109 (0.022-0.305) | 0.079 (0.000-0.169) | 0.278 (0.000-0.600) | 0.896 ( 0.873- 0.920) | 0.049 (0.000-0.106) |

#### 1.7 Table S7. Performance on FFR on the internal test set using the estimated stenosis

| Segments | CAGs | MAE | r | ROC AUC | PR AUC | F1 | Sensitivity | Specificity | Precision |
| --- | --- | --- | --- | --- | --- | --- | --- | --- | --- |
| 1 Proximal RCA | 57 | 0.164 (0.122-0.000) | 0.328 (0.084-0.000) | 0.638 (0.363-0.000) | 0.328 (0.084-0.555) | 0.249 (0.000-0.545) | 0.404 (0.000-1.000) | 0.846 (0.745-0.942) | 0.192 (0.000-0.455) |
| 2 Middle RCA | 101 | 0.252 (0.214-0.292) | 0.197 (-0.027-0.395) | 0.513 (0.366-0.684) | 0.197 (-0.027-0.395) | 0.258 (0.109-0.392) | 0.438 (0.190-0.667) | 0.579 (0.472-0.680) | 0.186 (0.079-0.300) |
| 3 Distale RCA | 41 | 0.238 (0.184-0.000) | 0.001 (-0.261-0.000) | 0.545 (0.344-0.000) | 0.001 (-0.261-0.288) | 0.308 (0.080-0.512) | 0.424 (0.154-0.750) | 0.484 (0.310-0.667) | 0.253 (0.071-0.467) |
| 4 PDA | 21 | 0.179 (0.101-0.000) | -0.224 (-0.564-0.000) | 0.500 (0.211-0.000) | -0.224 (-0.564-0.191) | 0.203 (0.000-0.571) | 0.197 (0.000-0.667) | 0.811 (0.600-1.000) | 0.255 (0.000-1.000) |
| 5 LM | 65 | 0.094 (0.077-0.000) | 0.227 (-0.016-0.000) |  |  |  |  |  |  |
| 6 Proximal LAD | 201 | 0.171 (0.154-0.000) | 0.336 (0.220-0.000) | 0.715 (0.637-0.000) | 0.336 (0.220-0.446) | 0.553 (0.458-0.641) | 0.568 (0.457-0.681) | 0.707 (0.629-0.784) | 0.538 (0.427-0.643) |
| 7 Middle LAD | 320 | 0.166 (0.152-0.000) | 0.288 (0.185-0.000) | 0.651 (0.591-0.000) | 0.288 (0.185-0.387) | 0.539 (0.464-0.611) | 0.532 (0.447-0.616) | 0.684 (0.617-0.747) | 0.546 (0.464-0.629) |
| 8 Distal LAD | 45 | 0.141 (0.111-0.000) | 0.274 (0.109-0.000) | 0.646 (0.483-0.000) | 0.274 (0.109-0.473) | 0.136 (0.000-0.316) | 0.075 (0.000-0.185) | 1.000 (1.000-1.000) | 0.879 (0.000-1.000) |
| 9 Diagonal 1 | 44 | 0.111 (0.071-0.000) | -0.192 (-0.432-0.000) | 0.466 (0.292-0.000) | -0.192 (-0.432-0.196) | 0.000 (0.000-0.000) | 0.000 (0.000-0.000) | 0.905 (0.800-1.000) | 0.000 (0.000-0.000) |
| 10 Diagonal 2 | 13 | 0.159 (0.090-0.000) | 0.014 (-0.556-0.000) | 0.611 (0.267-0.000) | 0.014 (-0.556-0.667) | 0.257 (0.000-0.667) | 0.250 (0.000-0.800) | 0.779 (0.500-1.000) | 0.321 (0.000-1.000) |
| 11 Proximal LCX | 80 | 0.108 (0.088-0.000) | 0.352 (0.176-0.000) | 0.624 (0.413-0.000) | 0.352 (0.176-0.546) | 0.255 (0.000-0.571) | 0.284 (0.000-0.667) | 0.917 (0.849-0.973) | 0.254 (0.000-0.616) |
| 12 Marginal 1 | 71 | 0.135 (0.108-0.000) | 0.154 (-0.013-0.000) | 0.587 (0.428-0.000) | 0.154 (-0.013-0.320) | 0.000 (0.000-0.000) | 0.000 (0.000-0.000) | 0.878 (0.791-0.955) | 0.000 (0.000-0.000) |
| 13 Middle LCX | 69 | 0.091 (0.068-0.000) | 0.147 (-0.038-0.000) | 0.589 (0.441-0.000) | 0.147 (-0.038-0.359) | 0.000 (0.000-0.000) | 0.000 (0.000-0.000) | 0.949 (0.885-1.000) | 0.000 (0.000-0.000) |
| 14 Marginal 2 | 22 | 0.123 (0.074-0.000) | -0.303 (-0.636-0.000) | 0.552 (0.250-0.000) | -0.303 (-0.636-0.343) | 0.000 (0.000-0.000) | 0.000 (0.000-0.000) | 0.872 (0.688-1.000) | 0.000 (0.000-0.000) |
| 15 Distal LCX | 19 | 0.112 (0.075-0.000) | 0.112 (-0.212-0.000) | 0.614 (0.316-0.000) | 0.112 (-0.212-0.507) | 0.000 (0.000-0.000) | 0.000 (0.000-0.000) | 0.927 (0.769-1.000) | 0.000 (0.000-0.000) |
| 16 PLA | 11 | 0.088 (0.058-0.000) | 0.344 (-0.316-0.000) | 0.700 (0.400-0.000) | 0.344 (-0.316-0.759) | 0.000 (0.000-0.000) | 0.000 (0.000-0.000) | 1.000 (1.000-1.000) | 0.000 (0.000-0.000) |

#### 1.8 Table S8. Performance on FFR on the internal test set using visual assessment baseline

| Segments | CAGs | MAE | r | ROC AUC | PR AUC | F1 | Sensitivity | Specificity | Precision |
| --- | --- | --- | --- | --- | --- | --- | --- | --- | --- |
| 1 Proximal RCA | 60 | 0.385 (0.359-0.410) | 0.616 (0.442-0.748) | 0.918 (0.774-1.000) | 0.697 (0.301-1.000) | 0.826 (0.500-1.000) | 0.839 (0.500-1.000) | 0.982 (0.943-1.000) | 0.835 (0.500-1.000) |
| 2 Middle RCA | 106 | 0.400 (0.379-0.420) | 0.586 (0.454-0.708) | 0.904 (0.829-0.961) | 0.681 (0.496-0.830) | 0.691 (0.518-0.847) | 0.760 (0.560-0.941) | 0.895 (0.826-0.953) | 0.643 (0.438-0.833) |
| 3 Distale RCA | 42 | 0.409 (0.371-0.445) | 0.570 (0.342-0.735) | 0.903 (0.765-0.999) | 0.854 (0.662-0.995) | 0.795 (0.583-0.952) | 0.833 (0.600-1.000) | 0.900 (0.800-1.000) | 0.768 (0.500-1.000) |
| 4 PDA | 17 | 0.504 (0.402-0.626) | 0.222 (-0.330-0.704) | 0.635 (0.345-0.886) | 0.384 (0.124-0.733) | 0.488 (0.154-0.800) | 0.756 (0.200-1.000) | 0.612 (0.333-0.891) | 0.386 (0.091-0.717) |
| 5 LM | 65 | 0.340 (0.313-0.367) | 0.649 (0.501-0.781) |  |  |  |  |  |  |
| 6 Proximal LAD | 200 | 0.365 (0.350-0.381) | 0.715 (0.636-0.782) | 0.913 (0.874-0.947) | 0.853 (0.788-0.908) | 0.771 (0.691-0.844) | 0.681 (0.571-0.785) | 0.953 (0.915-0.984) | 0.898 (0.814-0.968) |
| 7 Middle LAD | 321 | 0.369 (0.355-0.381) | 0.679 (0.621-0.733) | 0.892 (0.855-0.925) | 0.833 (0.771-0.886) | 0.745 (0.682-0.803) | 0.646 (0.563-0.729) | 0.942 (0.908-0.973) | 0.887 (0.821-0.945) |
| 8 Distal LAD | 45 | 0.393 (0.346-0.441) | 0.621 (0.476-0.743) | 0.814 (0.674-0.926) | 0.879 (0.778-0.953) | 0.737 (0.585-0.863) | 0.629 (0.452-0.808) | 0.887 (0.720-1.000) | 0.894 (0.737-1.000) |
| 9 Diagonal 1 | 44 | 0.466 (0.418-0.517) | 0.246 (-0.066-0.562) | 0.806 (0.662-0.929) | 0.574 (0.328-0.824) | 0.613 (0.385-0.800) | 0.835 (0.571-1.000) | 0.685 (0.529-0.833) | 0.501 (0.286-0.714) |
| 10 Diagonal 2 | 13 | 0.487 (0.366-0.602) | 0.396 (-0.269-0.898) | 0.903 (0.667-1.000) | 0.762 (0.333-1.000) | 0.880 (0.571-1.000) | 1.000 (1.000-1.000) | 0.889 (0.632-1.000) | 0.806 (0.400-1.000) |
| 11 Proximal LCX | 80 | 0.416 (0.388-0.447) | 0.636 (0.502-0.747) | 0.956 (0.894-0.993) | 0.600 (0.271-0.893) | 0.471 (0.230-0.688) | 1.000 (1.000-1.000) | 0.796 (0.700-0.882) | 0.323 (0.120-0.526) |
| 12 Marginal 1 | 72 | 0.457 (0.424-0.492) | 0.449 (0.242-0.658) | 0.865 (0.741-0.957) | 0.315 (0.091-0.634) | 0.385 (0.133-0.621) | 0.835 (0.500-1.000) | 0.787 (0.691-0.879) | 0.259 (0.063-0.474) |
| 13 Middle LCX | 69 | 0.404 (0.368-0.441) | 0.209 (-0.045-0.509) | 0.637 (0.399-0.835) | 0.354 (0.126-0.632) | 0.349 (0.125-0.571) | 0.449 (0.125-0.750) | 0.792 (0.684-0.893) | 0.290 (0.091-0.530) |
| 14 Marginal 2 | 22 | 0.480 (0.418-0.550) | 0.298 (-0.195-0.769) | 0.813 (0.598-0.978) | 0.567 (0.262-0.941) | 0.600 (0.200-0.857) | 0.673 (0.250-1.000) | 0.819 (0.600-1.000) | 0.573 (0.143-1.000) |
| 15 Distal LCX | 19 | 0.422 (0.353-0.485) | 0.632 (0.362-0.794) | 0.886 (0.683-1.000) | 0.735 (0.299-1.000) | 0.604 (0.222-0.857) | 0.810 (0.398-1.000) | 0.712 (0.500-0.929) | 0.503 (0.164-0.859) |
| 16 PLA | 9 | 0.448 (0.377-0.523) | 0.000 (0.000-000) | 0.875 (0.714-1.000) | 0.481 (0.200-1.000) | 0.615 (0.314-1.000) | 1.000 (1.000-1.000) | 0.737 (0.375-1.000) | 0.491 (0.200-1.000) |

#### 1.9 Table S9. Performance on predicting FFR on external test set using the estimated stenosis

| Segments | CAGs | MAE | r | ROC AUC | PR AUC | F1 | Sensitivity | Specificity | Precision |
| --- | --- | --- | --- | --- | --- | --- | --- | --- | --- |
| 1 Proximal RCA | 20 | 0.174 (0.105-0.259) | 0.215 (-0.204-0.796) | 0.569 (0.053-1.000) | 0.475 (0.056-1.000) | 0.278 (0.000-0.726) | 0.354 (0.000-1.000) | 0.831 (0.625-1.000) | 0.260 (0.000-1.000) |
| 2 Middle RCA | 62 | 0.275 (0.223-0.333) | 0.505 (0.292-0.693) | 0.790 (0.634-0.915) | 0.527 (0.272-0.764) | 0.507 (0.316-0.683) | 0.842 (0.600-1.000) | 0.618 (0.481-0.756) | 0.368 (0.200-0.542) |
| 3 Distale RCA | 126 | 0.073 (0.057-0.092) | 0.464 (0.319-0.602) | 0.911 (0.832-0.980) | 0.216 (0.053-0.502) | 0.196 (0.000-0.571) | 0.322 (0.000-1.000) | 0.951 (0.904-0.984) | 0.143 (0.000-0.429) |
| 4 PDA | 11 | 0.149 (0.056-0.278) | -0.060 (-0.608-0.651) |  |  |  |  |  |  |
| 5 LM | 15 | 0.111 (0.078-0.148) | 0.658 (0.438-0.818) |  |  |  |  |  |  |
| 6 Proximal LAD | 116 | 0.146 (0.122-0.172) | 0.279 (0.088-0.474) | 0.641 (0.550-0.749) | 0.570 (0.437-0.715) | 0.471 (0.329-0.600) | 0.391 (0.244-0.531) | 0.821 (0.727-0.909) | 0.606 (0.432-0.778) |
| 7 Middle LAD | 153 | 0.125 (0.107-0.147) | 0.540 (0.433-0.637) | 0.810 (0.734-0.883) | 0.683 (0.545-0.823) | 0.604 (0.485-0.718) | 0.514 (0.380-0.647) | 0.912 (0.850-0.962) | 0.741 (0.593-0.882) |
| 8 Distal LAD | 63 | 0.087 (0.070-0.106) | 0.365 (0.233-0.515) | 0.762 (0.634-0.879) | 0.336 (0.162-0.551) | 0.000 (0.000-0.000) | 0.000 (0.000-0.000) | 1.000 (1.000-1.000) | 0.000 (0.000-0.000) |
| 9 Diagonal 1 | 10 | 0.089 (0.042-0.143) | 0.254 (-0.364-0.770) | 0.762 (0.417-1.000) | 0.912 (0.700-1.000) | 0.000 (0.000-0.000) | 0.000 (0.000-0.000) | 1.000 (1.000-1.000) | 0.000 (0.000-0.000) |
| 10 Diagonal 2 | 4 | 0.103 (0.039-0.196) |  |  |  |  |  |  |  |
| 11 Proximal LCX | 22 | 0.095 (0.059-0.134) | 0.108 (-0.343-0.572) |  |  |  |  |  |  |
| 12 Marginal 1 | 68 | 0.113 (0.085-0.146) | 0.342 (0.142-0.549) | 0.712 (0.446-0.955) | 0.148 (0.027-0.429) | 0.000 (0.000-0.000) | 0.000 (0.000-0.000) | 0.939 (0.877-0.985) | 0.000 (0.000-0.000) |
| 13 Middle LCX | 109 | 0.089 (0.075-0.107) | 0.335 (0.143-0.524) | 0.818 (0.550-0.991) | 0.243 (0.022-0.667) | 0.235 (0.000-0.667) | 0.345 (0.000-1.000) | 0.962 (0.923-0.991) | 0.206 (0.000-0.667) |
| 14 Marginal 2 | 22 | 0.062 (0.044-0.079) | 0.573 (0.237-0.850) |  |  |  |  |  |  |
| 15 Distal LCX | 9 | 0.064 (0.039-0.094) | 0.376 (-0.253-0.941) |  |  |  |  |  |  |
| 16 PLA | 10 | 0.068 (0.047-0.087) | 0.518 (0.094-0.840) |  |  |  |  |  |  |

#### 1.10 Table S10. Performance on predicting FFR on external test dataset using the visual assessment

| Segments | CAGs | MAE | r | ROC AUC | PR AUC | F1 | Sensitivity | Specificity | Precision |
| --- | --- | --- | --- | --- | --- | --- | --- | --- | --- |
| 1 Proximal RCA | 20 | 0.399 (0.354-0.450) | 0.616 (0.021-0.879) | 0.941 (0.750-1.000) | 0.812 (0.200-1.000) | 0.399 (0.354-0.450) | 0.676 (0.000-1.000) | 0.883 (0.706-1.000) | 0.509 (0.000-1.000) |
| 2 Middle RCA | 62 | 0.399 (0.359-0.437) | 0.600 (0.485-0.700) | 0.921 (0.837-0.983) | 0.711 (0.447-0.905) | 0.399 (0.359-0.437) | 0.923 (0.749-1.000) | 0.838 (0.735-0.936) | 0.598 (0.389-0.813) |
| 3 Distale RCA | 126 | 0.088 (0.068-0.109) | 0.604 (0.408-0.751) | 0.959 (0.886-1.000) | 0.558 (0.083-1.000) | 0.088 (0.068-0.109) | 0.334 (0.000-1.000) | 0.976 (0.950-1.000) | 0.271 (0.000-1.000) |
| 4 PDA | 11 | 0.390 (0.243-0.547) | 0.313 (-0.274-0.700) | 0.708 (0.222-1.000) | 0.646 (0.143-1.000) | 0.390 (0.243-0.547) | 0.667 (0.000-1.000) | 0.510 (0.143-0.857) | 0.340 (0.000-0.750) |
| 5 LM | 15 | 0.322 (0.273-0.366) | 0.630 (0.368-0.846) |  |  |  |  |  |  |
| 6 Proximal LAD | 117 | 0.367 (0.343-0.391) | 0.440 (0.243-0.595) | 0.792 (0.705-0.870) | 0.685 (0.544-0.807) | 0.367 (0.343-0.391) | 0.632 (0.500-0.773) | 0.807 (0.708-0.891) | 0.707 (0.568-0.837) |
| 7 Middle LAD | 154 | 0.299 (0.272-0.330) | 0.533 (0.433-0.621) | 0.791 (0.718-0.860) | 0.632 (0.492-0.755) | 0.299 (0.272-0.330) | 0.539 (0.404-0.674) | 0.853 (0.789-0.918) | 0.652 (0.512-0.792) |
| 8 Distal LAD | 63 | 0.189 (0.153-0.225) | 0.808 (0.735-0.867) | 0.919 (0.845-0.978) | 0.677 (0.389-0.887) | 0.189 (0.153-0.225) | 0.499 (0.181-0.818) | 0.925 (0.852-0.982) | 0.557 (0.250-0.875) |
| 9 Diagonal 1 | 10 | 0.406 (0.301-0.492) | 0.351 (-0.182-0.863) | 0.571 (0.194-0.920) | 0.828 (0.533-0.977) | 0.406 (0.301-0.492) | 0.709 (0.333-1.000) | 0.321 (0.000-1.000) | 0.715 (0.333-1.000) |
| 10 Diagonal 2 | 4 | 0.443 (0.220-0.558) | 0.000 (0.000-0.000) | 1.000 (1.000-1.000) | 1.000 (1.000-1.000) | 0.443 (0.220-0.558) | 1.000 (1.000-1.000) | 0.657 (0.000-1.000) | 0.670 (0.250-1.000) |
| 11 Proximal LCX | 23 | 0.334 (0.249-0.414) | 0.725 (0.573-0.838) | 0.908 (0.682-1.000) | 0.828 (0.412-1.000) | 0.334 (0.249-0.414) | 0.740 (0.000-1.000) | 0.895 (0.737-1.000) | 0.602 (0.000-1.000) |
| 12 Marginal 1 | 69 | 0.159 (0.112-0.207) | 0.669 (0.472-0.796) | 0.993 (0.970-1.000) | 0.779 (0.200-1.000) | 0.159 (0.112-0.207) | 1.000 (1.000-1.000) | 0.925 (0.866-0.985) | 0.334 (0.111-0.667) |
| 13 Middle LCX | 109 | 0.188 (0.146-0.232) | 0.521 (0.351-0.665) | 0.703 (0.329-0.986) | 0.147 (0.009-0.487) | 0.188 (0.146-0.232) | 0.357 (0.000-1.000) | 0.848 (0.778-0.913) | 0.065 (0.000-0.203) |
| 14 Marginal 2 | 22 | 0.259 (0.146-0.385) | 0.629 (0.310-0.872) | 0.632 (0.263-0.925) | 0.251 (0.045-0.584) | 0.259 (0.146-0.385) | 0.321 (0.000-1.000) | 0.736 (0.528-0.938) | 0.170 (0.000-0.500) |
| 15 Distal LCX | 9 | 0.147 (0.047-0.270) | 0.000 (0.000-0.000) |  |  |  |  |  |  |
| 16 PLA | 10 | 0.377 (0.201-0.544) | 0.624 (-0.025-0.947) | 0.444 (0.167-0.750) | 0.227 (0.111-0.500) | 0.377 (0.201-0.544) | 0.000 (0.000-0.000) | 0.562 (0.222-0.889) | 0.000 (0.000-0.000) |

#### 1.11 Table S11. Performance on predicting QCA on the external test dataset using estimated stenosis

| Segments | CAGs | MAE | r | ROC AUC | PR AUC | F1 | Sensitivity | Specificity | Precision |
| --- | --- | --- | --- | --- | --- | --- | --- | --- | --- |
| 1 Proximal RCA | 207 | 0.241 (0.222-0.260) | 0.648 (0.505-0.763) | 0.899 (0.795-0.970) | 0.695 (0.480-0.869) | 0.580 (0.375-0.750) | 0.668 (0.444-0.880) | 0.942 (0.904-0.973) | 0.527 (0.313-0.733) |
| 2 Middle RCA | 111 | 0.178 (0.158-0.199) | 0.668 (0.540-0.760) | 0.976 (0.940-1.000) | 0.670 (0.336-1.000) | 0.237 (0.087-0.391) | 1.000 (1.000-1.000) | 0.637 (0.538-0.727) | 0.139 (0.044-0.250) |
| 3 Distale RCA | 69 | 0.183 (0.156-0.212) | 0.611 (0.422-0.766) | 0.934 (0.851-1.000) | 0.454 (0.111-1.000) | 0.268 (0.091-0.500) | 1.000 (1.000-1.000) | 0.741 (0.632-0.848) | 0.160 (0.045-0.333) |
| 4 PDA | 46 | 0.331 (0.287-0.380) | 0.358 (0.102-0.615) | 0.888 (0.763-0.977) | 0.478 (0.179-0.862) | 0.361 (0.000-0.727) | 0.388 (0.000-1.000) | 0.927 (0.837-1.000) | 0.403 (0.000-1.000) |
| 5 LM | 50 | 0.170 (0.141-0.203) | 0.365 (0.064-0.604) |  |  |  |  |  |  |
| 6 Proximal LAD | 162 | 0.133 (0.119-0.150) | 0.546 (0.424-0.656) | 0.885 (0.799-0.968) | 0.269 (0.061-0.583) | 0.143 (0.047-0.253) | 1.000 (1.000-1.000) | 0.545 (0.468-0.622) | 0.077 (0.024-0.147) |
| 7 Middle LAD | 220 | 0.142 (0.128-0.157) | 0.570 (0.457-0.666) | 0.837 (0.701-0.949) | 0.521 (0.236-0.773) | 0.209 (0.103-0.323) | 0.771 (0.500-1.000) | 0.646 (0.580-0.708) | 0.120 (0.050-0.191) |
| 8 Distal LAD | 107 | 0.243 (0.220-0.269) | 0.271 (0.082-0.446) | 0.594 (0.512-0.689) | 0.035 (0.019-0.077) | 0.000 (0.000-0.000) | 0.000 (0.000-0.000) | 0.945 (0.896-0.981) | 0.000 (0.000-0.000) |
| 9 Diagonal 1 | 131 | 0.271 (0.249-0.293) | 0.313 (0.076-0.523) | 0.721 (0.248-1.000) | 0.398 (0.032-1.000) | 0.232 (0.000-0.667) | 0.253 (0.000-1.000) | 0.977 (0.951-1.000) | 0.260 (0.000-1.000) |
| 10 Diagonal 2 | 25 | 0.298 (0.241-0.357) | -0.014 (-0.373-0.365) |  |  |  |  |  |  |
| 11 Proximal LCX | 109 | 0.201 (0.182-0.223) | 0.555 (0.316-0.736) | 0.931 (0.808-1.000) | 0.613 (0.083-1.000) | 0.329 (0.000-0.609) | 0.755 (0.200-1.000) | 0.896 (0.833-0.951) | 0.224 (0.000-0.500) |
| 12 Marginal 1 | 74 | 0.227 (0.198-0.256) | 0.304 (0.093-0.504) | 0.549 (0.028-0.875) | 0.104 (0.014-0.253) | 0.000 (0.000-0.000) | 0.000 (0.000-0.000) | 0.873 (0.791-0.944) | 0.000 (0.000-0.000) |
| 13 Middle LCX | 86 | 0.236 (0.210-0.260) | 0.630 (0.408-0.777) | 0.869 (0.698-1.000) | 0.528 (0.082-1.000) | 0.341 (0.000-0.625) | 0.600 (0.000-1.000) | 0.888 (0.813-0.953) | 0.248 (0.000-0.500) |
| 14 Marginal 2 | 20 | 0.273 (0.215-0.331) | 0.567 (0.337-0.790) | 0.843 (0.641-1.000) | 0.507 (0.167-1.000) | 0.000 (0.000-0.000) | 0.000 (0.000-0.000) | 0.886 (0.723-1.000) | 0.000 (0.000-0.000) |
| 15 Distal LCX | 17 | 0.298 (0.231-0.371) | 0.256 (-0.145-0.612) | 0.688 (0.438-0.875) | 0.266 (0.111-0.571) | 0.000 (0.000-0.000) | 0.000 (0.000-0.000) | 0.939 (0.813-1.000) | 0.000 (0.000-0.000) |
| 16 PLA | 44 | 0.292 (0.247-0.337) | 0.465 (0.092-0.776) | 0.826 (0.631-0.977) | 0.510 (0.117-0.893) | 0.420 (0.000-0.800) | 0.390 (0.000-1.000) | 0.947 (0.868-1.000) | 0.503 (0.000-1.000) |

#### 1.12 Table S12. Performance on predicting QCA on external dataset using visual assessment

| Segments | CAGs | MAE | r | ROC AUC | PR AUC | F1 | Sensitivity | Specificity | Precision |
| --- | --- | --- | --- | --- | --- | --- | --- | --- | --- |
| 1 Proximal RCA | 209 | 0.334 (0.312-0.359) | 0.432 (0.231-0.597) | 0.694 (0.560-0.830) | 0.444 (0.215-0.660) | 0.490 (0.258-0.686) | 0.444 (0.200-0.692) | 0.969 (0.941-0.990) | 0.578 (0.300-0.875) |
| 2 Middle RCA | 112 | 0.293 (0.267-0.322) | 0.600 (0.407-0.749) | 0.862 (0.627-0.991) | 0.519 (0.093-0.861) | 0.321 (0.095-0.545) | 0.834 (0.500-1.000) | 0.822 (0.748-0.895) | 0.205 (0.054-0.389) |
| 3 Distale RCA | 69 | 0.350 (0.324-0.379) | 0.576 (0.306-0.761) | 0.811 (0.434-1.000) | 0.548 (0.014-1.000) | 0.428 (0.000-0.800) | 0.660 (0.000-1.000) | 0.940 (0.876-0.985) | 0.339 (0.000-0.763) |
| 4 PDA | 46 | 0.410 (0.354-0.472) | 0.272 (-0.038-0.588) | 0.627 (0.366-0.984) | 0.384 (0.043-0.833) | 0.276 (0.000-0.588) | 0.401 (0.000-1.000) | 0.828 (0.702-0.946) | 0.222 (0.000-0.545) |
| 5 LM | 50 | 0.304 (0.271-0.341) | 0.388 (0.041-0.645) | 0.659 (0.372-0.969) | 0.355 (0.080-0.707) | 0.275 (0.000-0.522) | 0.495 (0.000-1.000) | 0.729 (0.595-0.845) | 0.206 (0.000-0.429) |
| 6 Proximal LAD | 162 | 0.323 (0.300-0.346) | 0.346 (0.194-0.492) | 0.735 (0.398-0.997) | 0.440 (0.035-0.857) | 0.129 (0.033-0.250) | 0.668 (0.200-1.000) | 0.672 (0.597-0.741) | 0.072 (0.016-0.153) |
| 7 Middle LAD | 220 | 0.338 (0.313-0.362) | 0.308 (0.165-0.447) | 0.542 (0.378-0.715) | 0.148 (0.044-0.318) | 0.160 (0.042-0.308) | 0.315 (0.077-0.600) | 0.843 (0.796-0.891) | 0.113 (0.026-0.222) |
| 8 Distal LAD | 107 | 0.369 (0.347-0.393) | 0.285 (0.101-0.458) | 0.458 (0.429-0.485) | 0.015 (0.009-0.037) | 0.000 (0.000-0.000) | 0.000 (0.000-0.000) | 0.943 (0.895-0.981) | 0.000 (0.000-0.000) |
| 9 Diagonal 1 | 131 | 0.422 (0.396-0.448) | 0.170 (0.054-0.296) | 0.457 (0.430-0.480) | 0.031 (0.008-0.061) | 0.000 (0.000-0.000) | 0.000 (0.000-0.000) | 0.930 (0.883-0.972) | 0.000 (0.000-0.000) |
| 10 Diagonal 2 | 25 | 0.402 (0.348-0.450) |  |  |  |  |  |  |  |
| 11 Proximal LCX | 109 | 0.330 (0.302-0.356) | 0.563 (0.383-0.701) | 0.690 (0.379-0.996) | 0.335 (0.009-0.917) | 0.204 (0.000-0.462) | 0.499 (0.000-1.000) | 0.875 (0.814-0.934) | 0.136 (0.000-0.333) |
| 12 Marginal 1 | 74 | 0.366 (0.339-0.394) | 0.517 (0.330-0.680) | 0.723 (0.384-0.959) | 0.164 (0.014-0.458) | 0.139 (0.000-0.400) | 0.325 (0.000-1.000) | 0.859 (0.771-0.931) | 0.094 (0.000-0.300) |
| 13 Middle LCX | 86 | 0.388 (0.352-0.422) | 0.265 (0.029-0.502) | 0.622 (0.369-0.931) | 0.240 (0.023-0.653) | 0.194 (0.000-0.437) | 0.416 (0.000-1.000) | 0.838 (0.749-0.925) | 0.133 (0.000-0.333) |
| 14 Marginal 2 | 20 | 0.359 (0.297-0.420) |  | 1.000 (1.000-1.000) | 1.000 (1.000-1.000) | 1.000 (1.000-1.000) | 1.000 (1.000-1.000) | 1.000 (1.000-1.000) | 1.000 (1.000-1.000) |
| 15 Distal LCX | 17 | 0.444 (0.380-0.513) |  |  |  |  |  |  |  |
| 16 PLA | 44 | 0.389 (0.338-0.443) | 0.323 (-0.008-0.633) | 0.538 (0.380-0.795) | 0.284 (0.045-0.689) | 0.203 (0.000-0.571) | 0.209 (0.000-0.667) | 0.924 (0.829-1.000) | 0.230 (0.000-1.000) |

### 2. Supplemental Figures

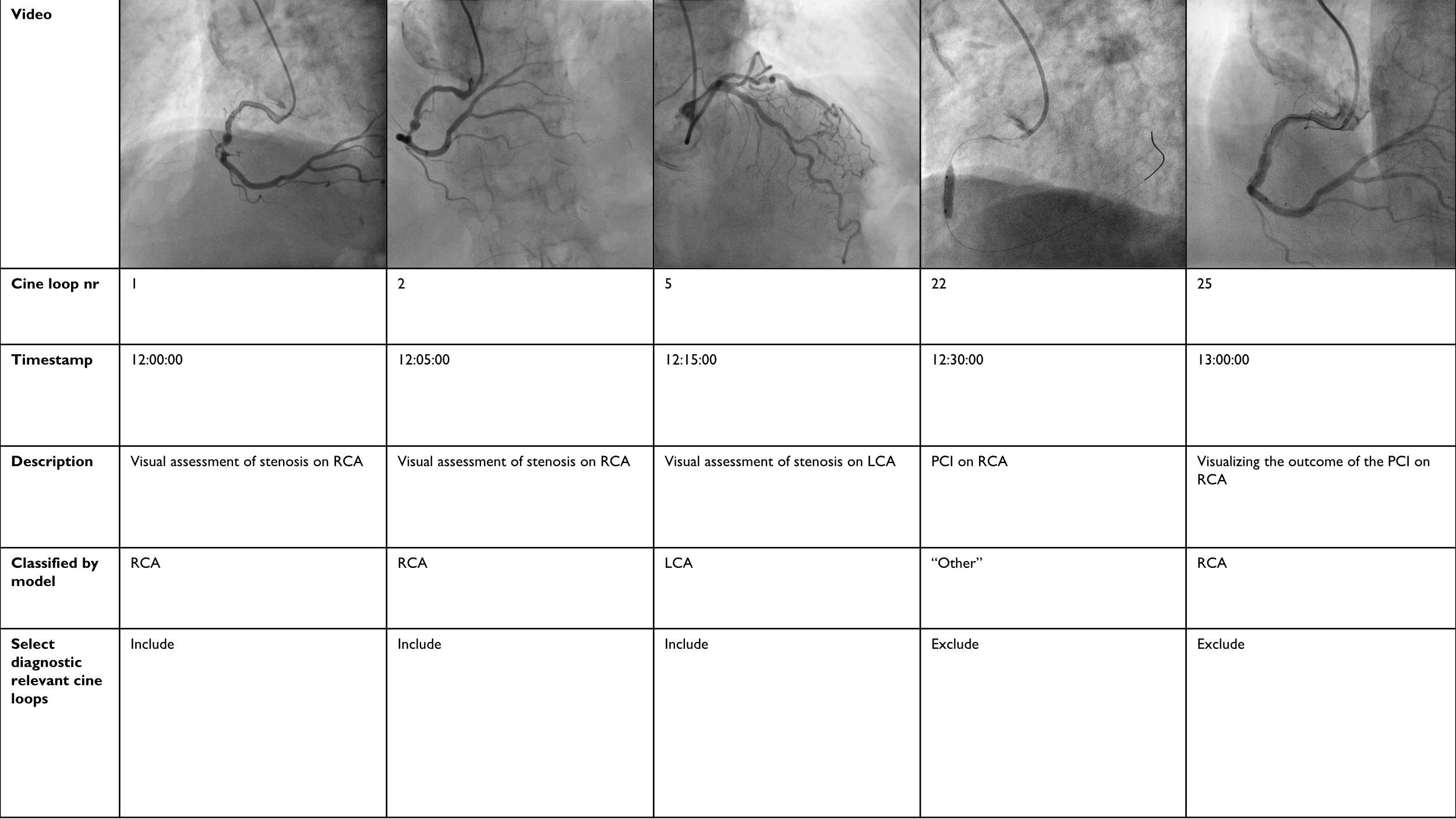

#### Figure S1. Inclusion of cine loops

#### The inclusion of cine loops is carried out by selecting all diagnostically relevant cine loops for the left and right coronary artery individually. For the RCA, this involves selecting all cine loops in a CAG with timestamps before the “other” category appears, indicating that a PCI has been initiated. The same procedure is carried out for the LCA. This is implemented with database operations (queries), aiming to remove cine loops during and after PCI.

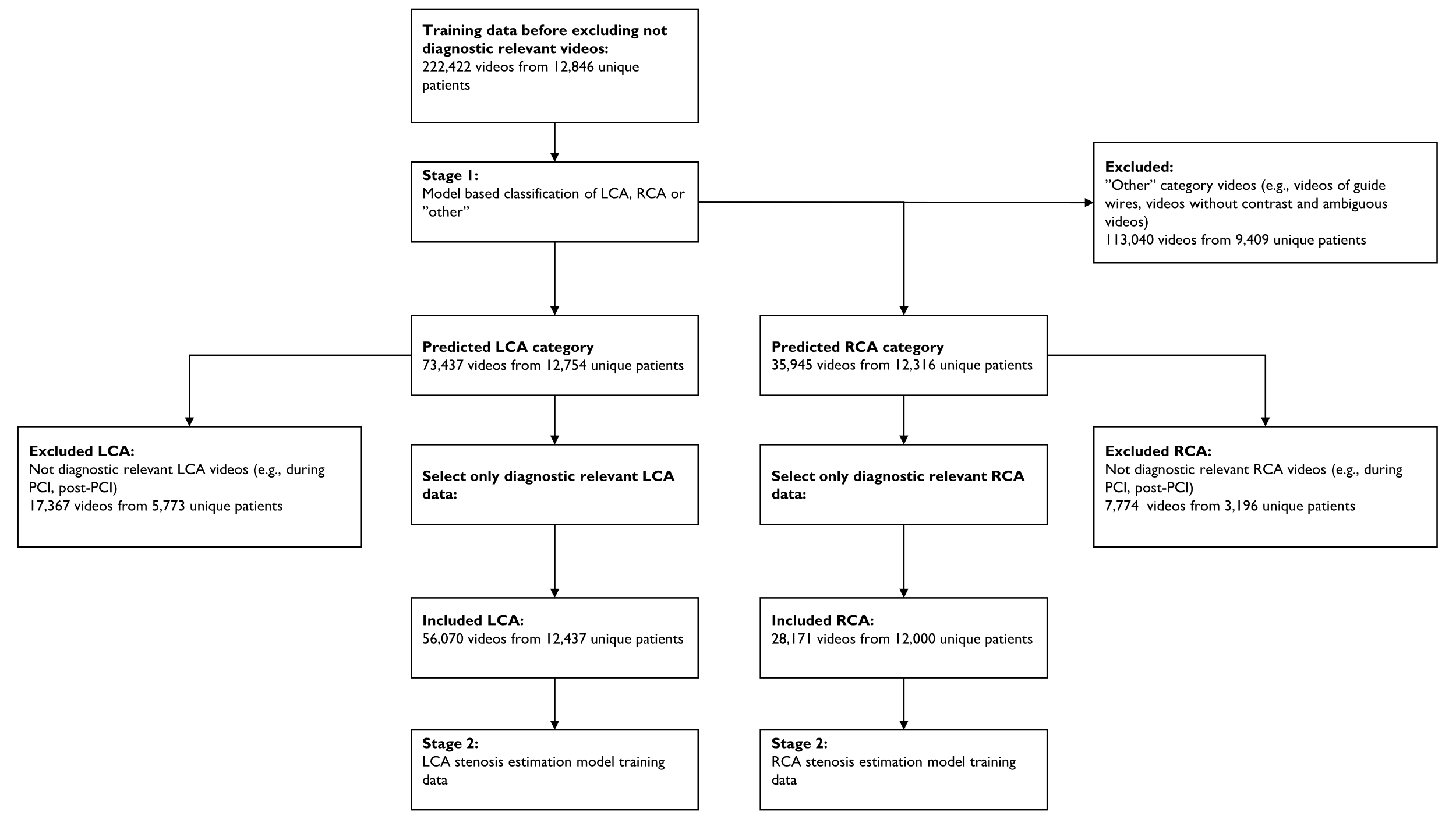

#### Figure S2: Flowchart for inclusion of cine loops in the training set

The flowchart describes how diagnostically relevant cine loops (videos) are included in the training dataset. As exclusion is performed at the cine loop level, the flowchart concerns cine loops and not patients. Occasionally, the model classifies all cine loops in a CAG as “other,” resulting in the complete exclusion of some CAGs (409 patients for the LCA and 846 for the RCA).

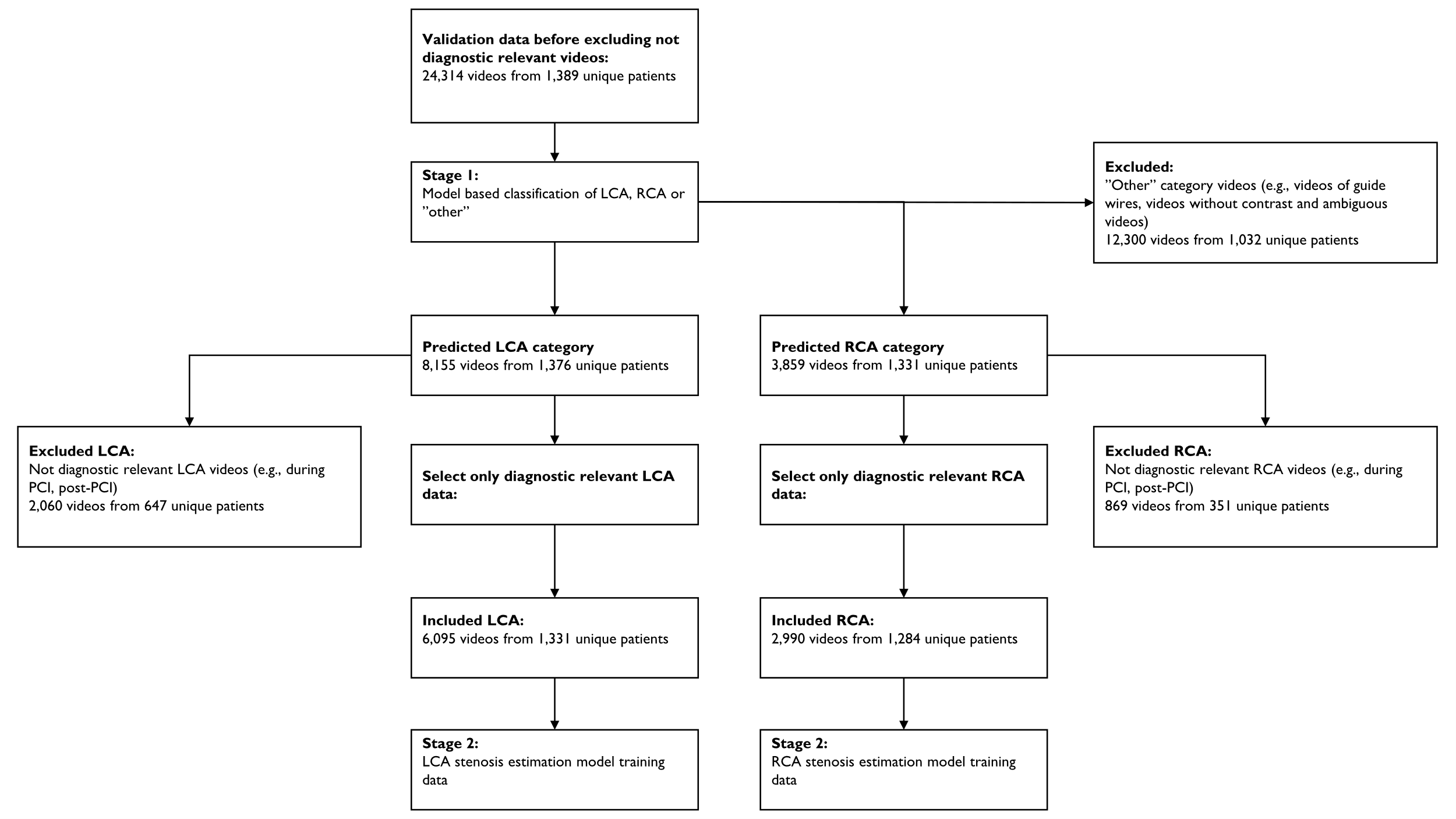

#### Figure S3: Flowchart for inclusion of cine loops in the validation set

The flowchart describes how diagnostically relevant cine loops (videos) are included in the validation dataset. As exclusion is performed at the cine loop level, the flowchart concerns cine loops and not patients. Occasionally, the model classifies all cine loops in a CAG as “other,” resulting in the complete exclusion of some CAGs (58 patients for the LCA and 105 for the RCA).

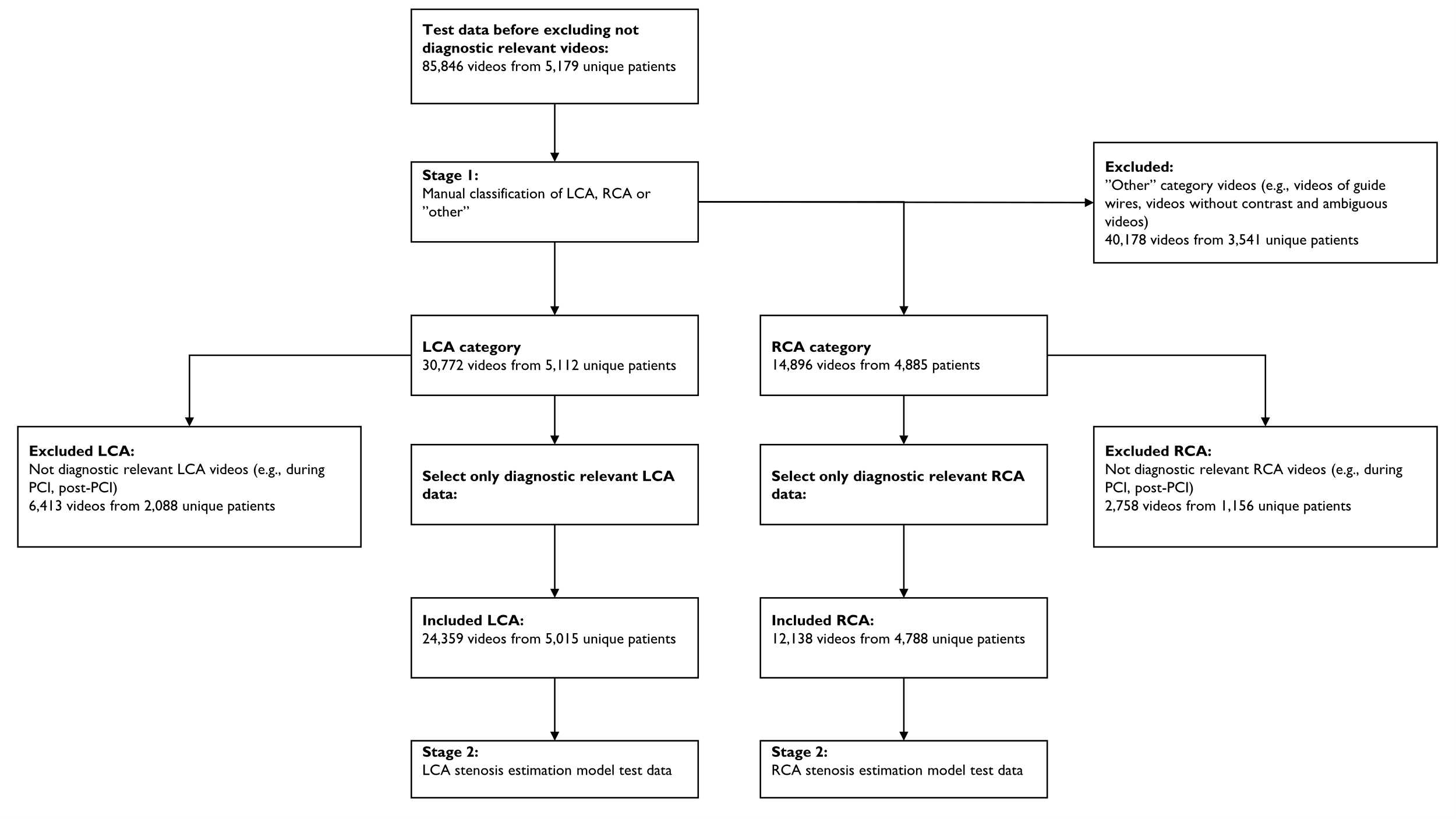

#### Figure S4: Flowchart for inclusion of cine loops in the internal test set

The flowchart describes how diagnostically relevant cine loops (videos) are included in the test dataset. As exclusion is performed at the cine loop level, the flowchart concerns cine loops and not patients. Occasionally, the model classifies all cine loops in a CAG as “other,” resulting in the complete exclusion of some CAGs (164 patients for the LCA and 391 for the RCA).

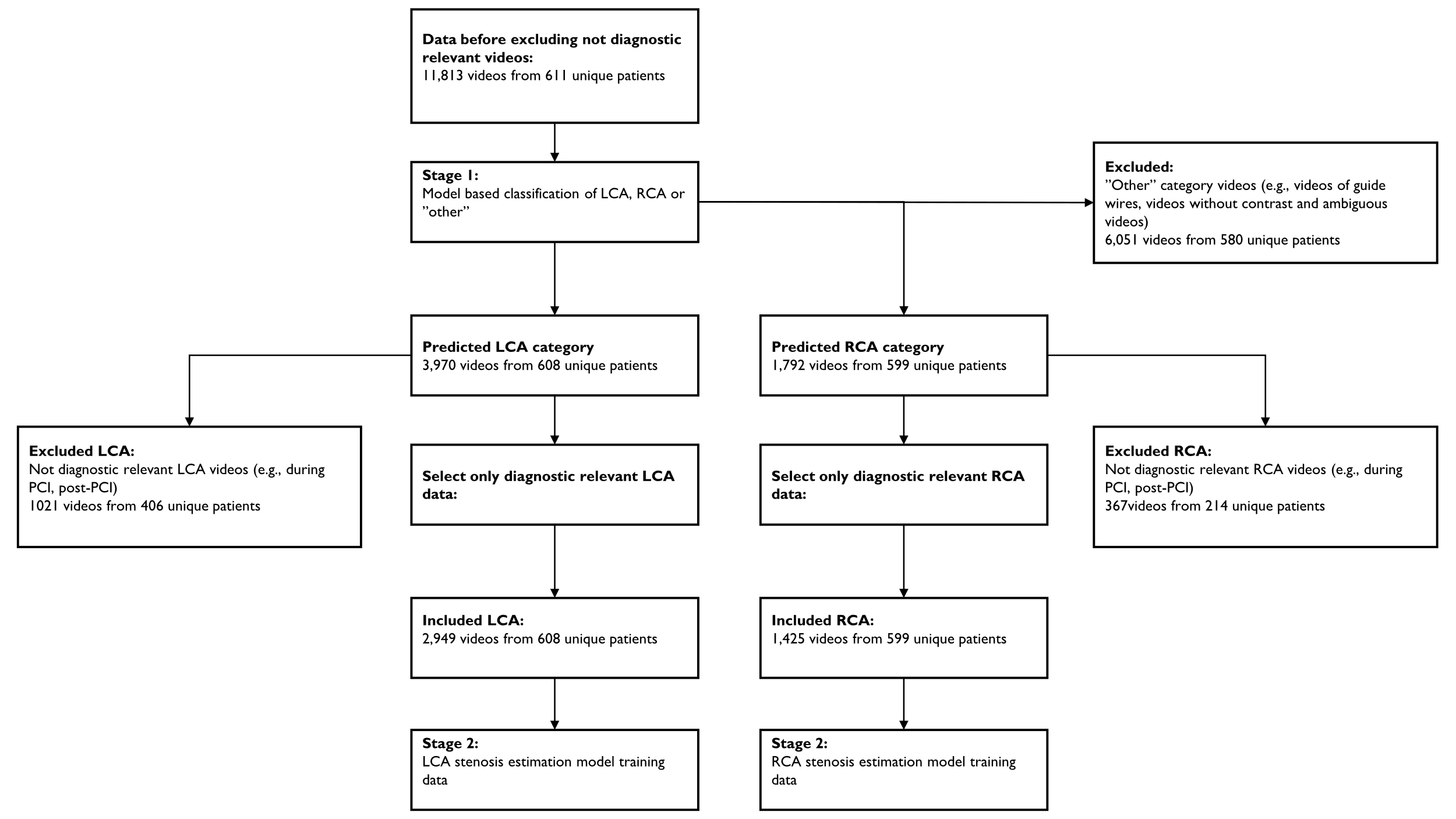

#### Figure S5: Flowchart for inclusion of cine loops in the external test dataset

The flowchart describes how diagnostically relevant cine loops (videos) are included in the external test dataset. As exclusion is performed at the cine loop level, the flowchart concerns cine loops and not patients. Occasionally, the model classifies all cine loops in a CAG as “other,” resulting in the complete exclusion of some CAGs (3 patients for the LCA and 12 for the RCA).

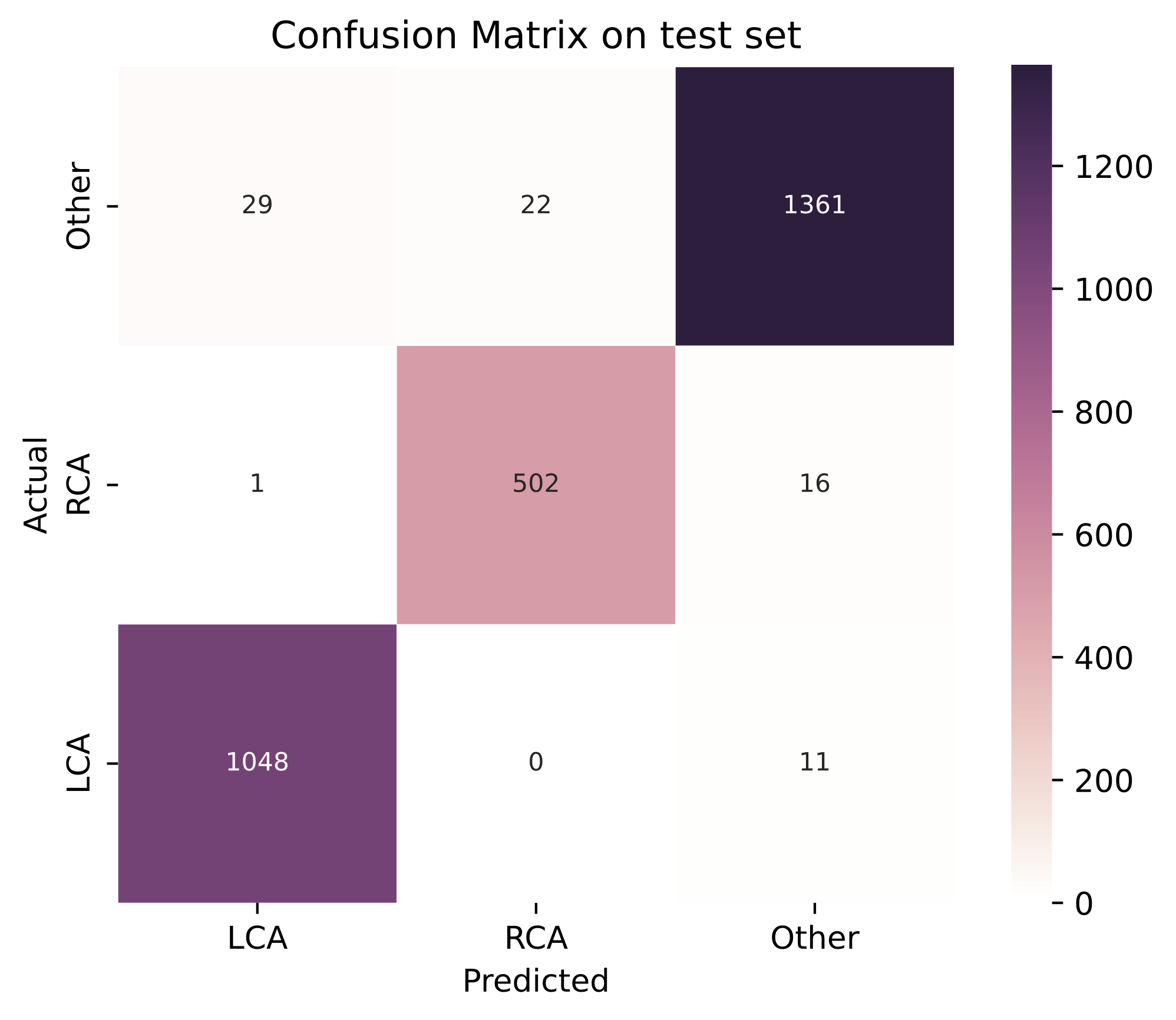

#### Figure S6: Confusion matrix for the cine loop classification model

The confusion matrix shows the performance of the cine loop classification model in stage 1. The rows in the confusion matrix are the actual categories and the columns are the actual categories (the manually labeled videos). The confusion matrix shows that we in total have 79 mispredictions (the off diagonal) for the test set with 2,990 videos form 179 patients.

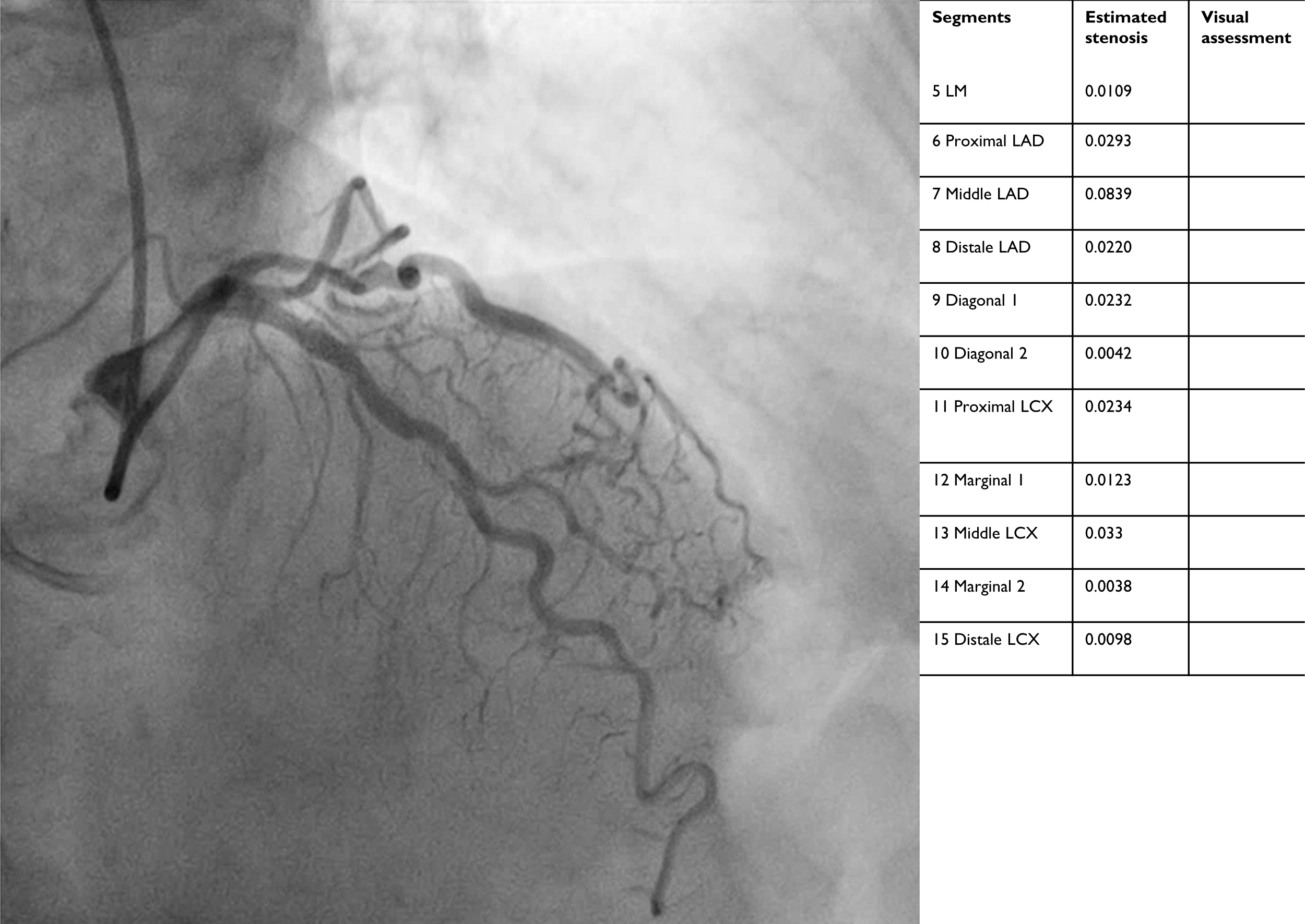

#### Figure S7. Static example of left coronary artery and the estimated stenoses

This figure presents a static example of the left coronary artery and the estimated stenoses. Notably, the output of the stenosis estimation model includes all segments, while the manual assessment does not, as the cardiologist found no significant stenosis. The stenosis estimation models did not detect any significant stenosis; however, they still attempt to quantify stenosis in all segments. For example, the model estimates 8% stenosis in the middle of the LAD.

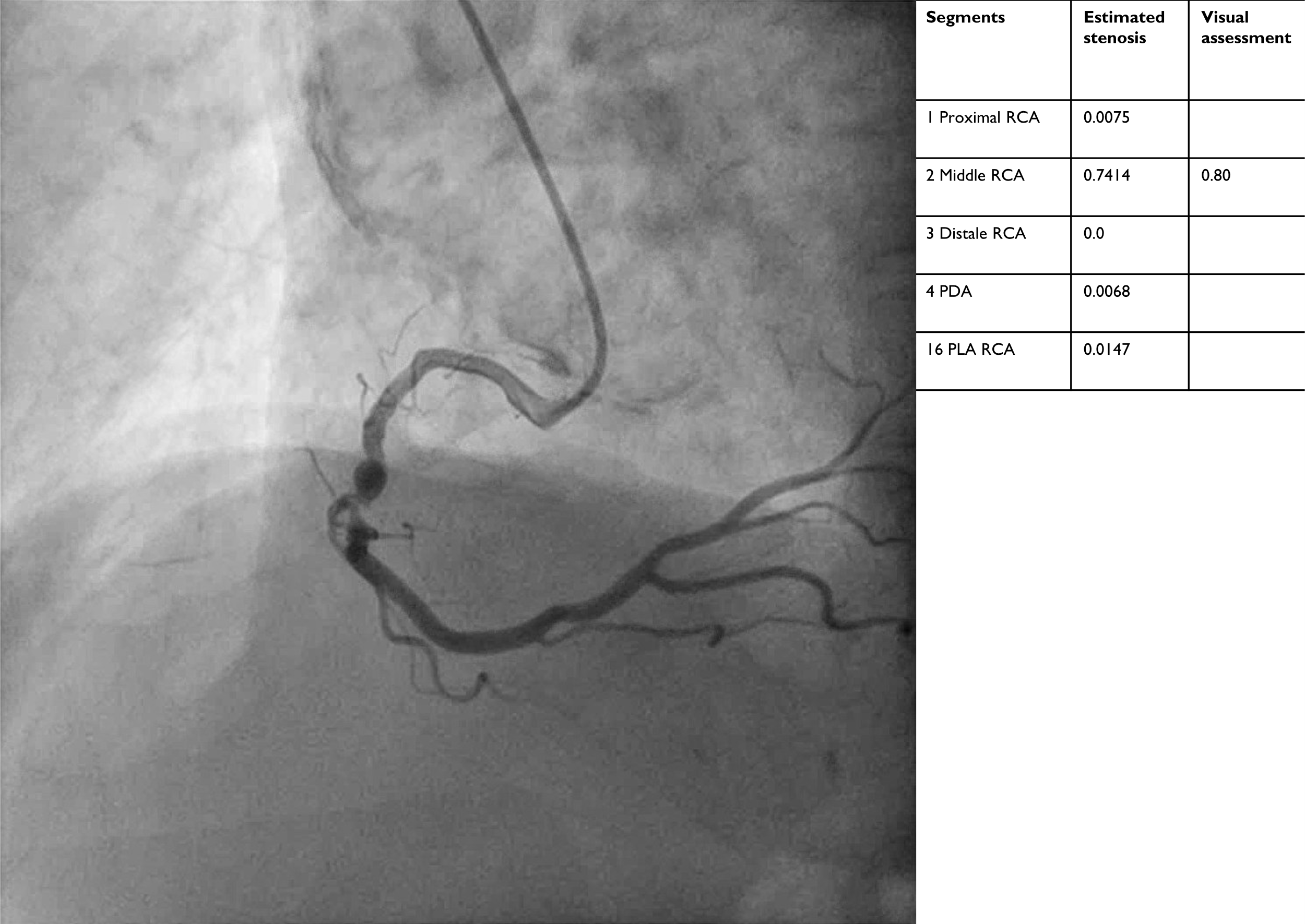

#### Figure S8. Static example of right coronary artery and the estimated stenoses

This figure shows a static example of the right coronary artery and the estimated stenoses. Notably, the stenosis estimation model outputs a 74% stenosis, aligning with the cardiologist’s report of 74% stenosis in the middle of the RCA.

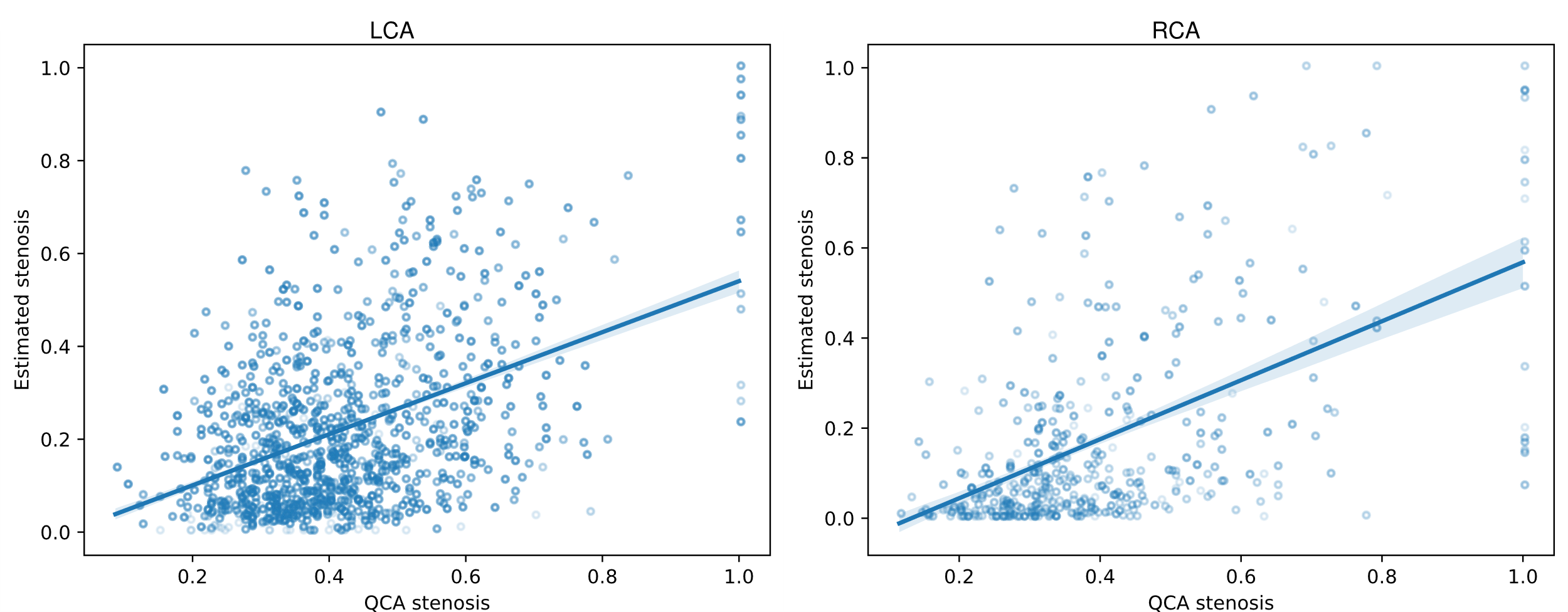

Figure S9. External dataset: Scatter plot of QCA and estimated stenosis

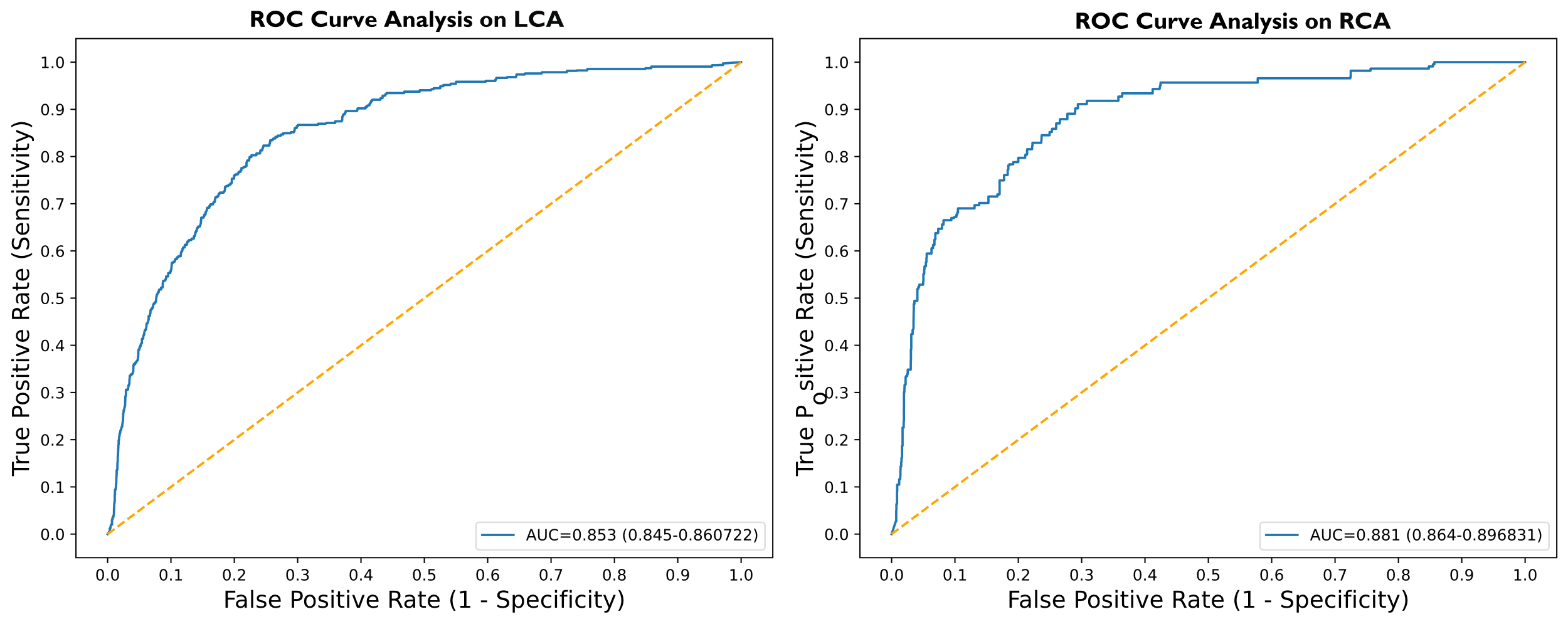

Figure S10: External dataset: Performance on detecting significant stenoses using visual assessment as ground truth

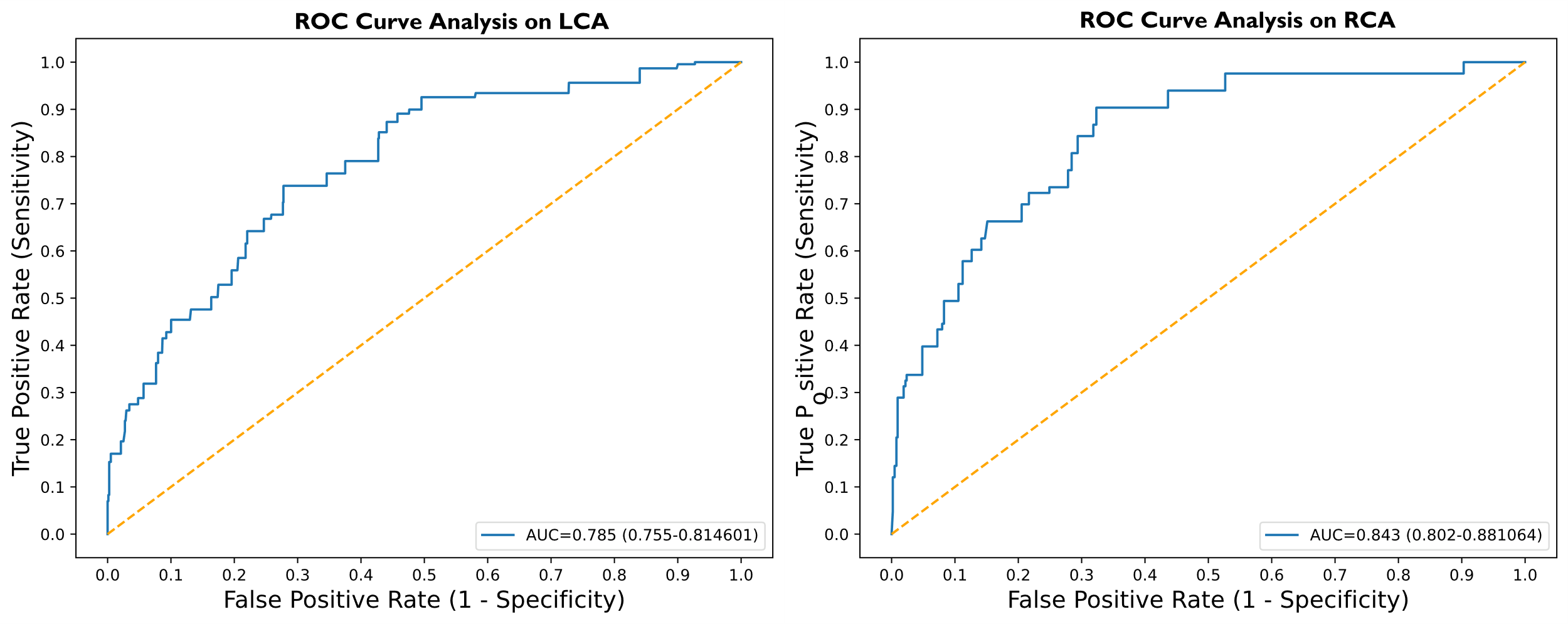

Figure S11: External dataset: Performance on detecting significant stenoses using QCA as ground truth

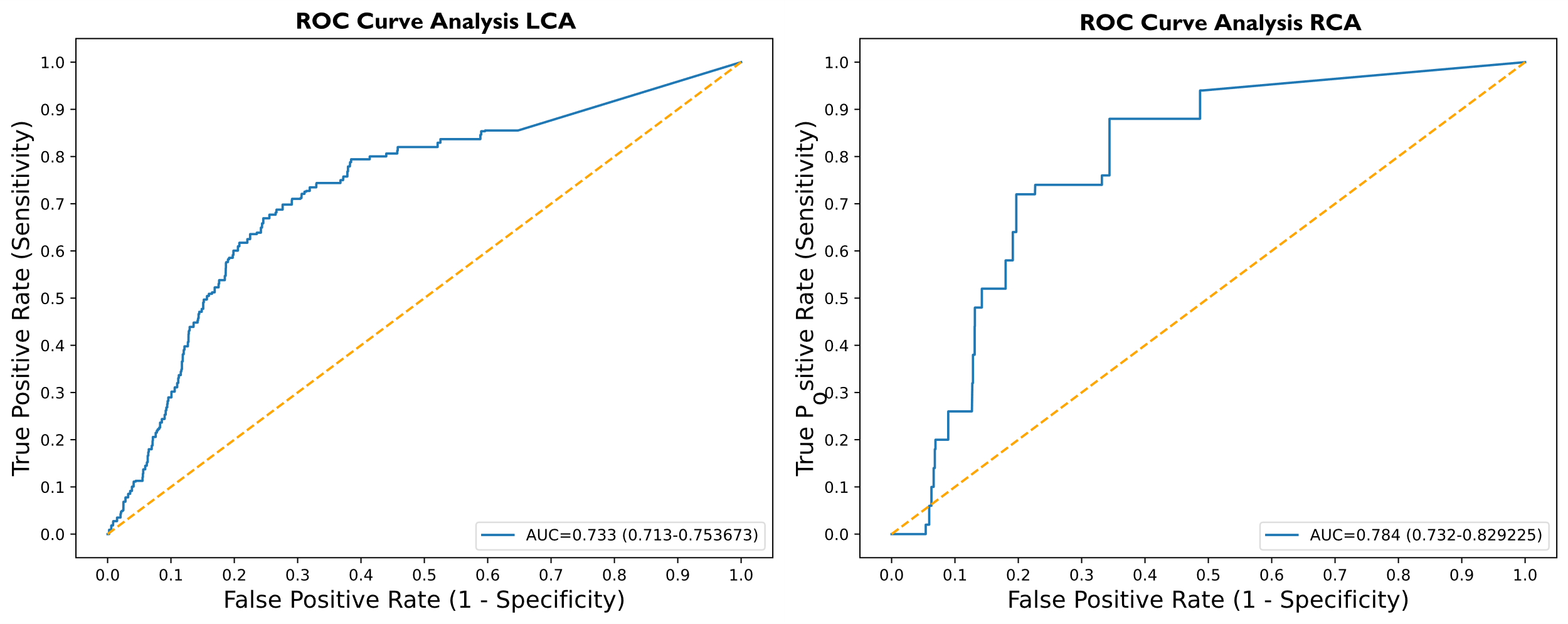

Figure S12: External dataset: Performance on detecting significant stenoses using FFR as ground truth

### 3. Supplemental videos legends

#### Video S1. Dynamic example of left coronary artery and the estimated stenoses

This video presents a dynamic example of the left coronary artery and the estimated stenoses. Notably, the output of the stenosis estimation model includes all segments, while the manual assessment does not, as the cardiologist found no significant stenosis. The stenosis estimation models did not detect any significant stenosis; however, they still attempt to quantify stenosis in all segments. For example, the model estimates 8% stenosis in the middle of the LAD.

#### Video S2. Dynamic example of right coronary artery and the estimated stenoses

This video shows a dynamic example of the right coronary artery and the estimated stenoses. Notably, the stenosis estimation model outputs a 74% stenosis, aligning with the cardiologist’s report of 74% stenosis in the middle of the RCA.
